# Pre-referral rectal artesunate and referral completion among children with suspected severe malaria in the Democratic Republic of the Congo, Nigeria and Uganda

**DOI:** 10.1101/2021.09.27.21264073

**Authors:** Nina C. Brunner, Elizabeth Omoluabi, Phyllis Awor, Jean Okitawutshu, Antoinette Tshefu, Aita Signorell, Babatunde Akano, Kazeem Ayodeji, Charles Okon, Ocheche Yusuf, Proscovia Athieno, Joseph Kimera, Gloria Tumukunde, Irene Angiro, Jean-Claude Kalenga, Giulia Delvento, Tristan T. Lee, Mark Lambiris, Amanda Ross, Nadja Cereghetti, Theodoor Visser, Harriet G. Napier, Valentina Buj, Christian Burri, Christian Lengeler, Manuel W. Hetzel

## Abstract

**Background:** Children who receive pre-referral rectal artesunate (RAS) require urgent referral to a health facility where appropriate treatment for severe malaria can be provided. However, the rapid improvement of a child’s condition after RAS administration may influence a caregiver’s decision to follow this recommendation. Currently, the evidence on the effect of RAS on referral completion is limited. In this study, we investigated the relationship between RAS implementation and administration and referral completion.

**Methods and Findings:** An observational study accompanied the roll-out of RAS in three malaria endemic settings in the Democratic Republic of the Congo (DRC), Nigeria and Uganda. Community health workers and primary health centres enrolled children under five years with suspected severe malaria before and after the roll-out of RAS. All children were followed up 28 days after enrolment to assess their treatment seeking pathways, treatments received, and their health outcome. In total, 8,365 children were enrolled, 77% of whom fulfilled all inclusion criteria and had a known referral completion status. Referral completion was 67% (1,408/2,104) in DRC, 48% (287/600) in Nigeria and 58% (2,170/3,745) in Uganda. In DRC and Uganda, RAS users were less likely to complete referral than RAS non-users in the pre-roll-out phase (adjusted odds ratio [aOR] = 0.48, 95% CI 0.30–0.77 and aOR = 0.72, 95% CI 0.58–0.88, respectively). Among children seeking care from a primary health centre in Nigeria, RAS users were less likely to complete referral compared to RAS non-users in the post-roll-out phase (aOR = 0.18, 95% CI 0.05–0.71). In Uganda, among children who completed referral, RAS users were significantly more likely to complete referral on time than RAS non-users enrolled in the pre-roll-out phase (aOR = 1.81, 95% CI 1.17–2.79).

**Conclusions:** The findings of this study raise legitimate concerns that the roll-out of RAS may lead to lower referral completion in children who were administered pre-referral RAS. To ensure that community-based programmes are effectively implemented, barriers to referral completion need to be addressed at all levels. Alternative effective treatment options should be provided to children unable to complete referral.

## Introduction

Rectal artesunate (RAS) is a potentially life-saving pre-referral treatment for children presenting at the primary health care level with suspected severe malaria [1]. Current guidelines require that children who received RAS be referred immediately to a health facility where comprehensive management of severe malaria can be provided [2]. However, the rapid improvement of a child’s condition after the administration of RAS [3] may result in children not being taken to a referral health facility (RHF) where appropriate treatment is available [4].

According to World Health Organization (WHO) guidelines, appropriate post-referral treatment of severe malaria consists of an intramuscular or intravenous antimalarial for at least 24 hours followed by a full course of an oral artemisinin-based combination therapy (ACT) and accompanied by the management of clinical complications [5]. Previous studies on RAS and referral defined referral completion as going to the nearest health facility, irrespective of the facility’s capacity to treat severe malaria [4, 6-10]. In the case of children first treated with RAS by a community health worker (CHW), this may be a primary health centre (PHC) that lacks the capacity to manage a severe malaria episode. In view of improving the case management of such children, more evidence is needed to understand the pathways by which children with suspected severe malaria reach a competent and capacitated health care provider and whether referral completion is impacted by the administration of RAS.

There is evidence that children who received pre-referral treatment were less likely to complete referral than children without treatment prior to referral [11, 12]. However, this kind of evidence for RAS as a pre-referral treatment is scarce. Most previous quantitative studies on RAS and referral completion did not compare RAS users versus non-RAS users [8-11, 13], and thus did not estimate the potential effect of RAS administration on referral completion. Only one observational study tested for non-inferiority of referral completion among children receiving RAS compared to children not receiving RAS [6]. The authors concluded non-inferiority because the pre-defined margin of 15% was not reached; however, referral completion in RAS users (84%) was lower than in non-RAS users (94%). In addition, the analysis did not control for other factors influencing referral completion. Factors that have previously been shown to influence referral completion are household dynamics and priorities, illness severity, the type of referring provider, the performance and result of diagnostic tests prior to referral, health workers’ communication skills, distance to the RHF, referral and treatment costs, and the perceived quality of the RHF [4, 6-8, 11-16].

The Community Access to Rectal Artesunate for Malaria (CARAMAL) project included an observational study accompanying the implementation of RAS in the Democratic Republic of the Congo (DRC), Nigeria and Uganda and is described in detail in a companion paper [17]. The training provided to CHW and staff of PHCs during the roll-out of RAS emphasised the need to refer severely sick children to an appropriate and recognised RHF capable of managing the child’s severe condition, rather than to the nearest or next-higher level provider. This manuscript aimed to assess referral completion of children with suspected severe malaria and its relationship with RAS implementation and administration, taking into consideration other factors influencing referral completion. The key study results are described elsewhere [18] and the treatment patterns in the RHF are described in [19].

## Method

### Study design

This observational study followed the implementation of pre-referral RAS in three study areas in DRC, Nigeria and Uganda. Local health authorities in collaboration with UNICEF trained community-based providers on the use and administration of rectal artesunate. Community-based providers enrolled children under five years of age with fever and danger signs according to the national integrated community case management (iCCM, in the case of CHW) or integrated management of childhood illness (IMCI, in the case of PHC) guidelines. All eligible children were followed up one month after enrolment by dedicated research staff.

The study period covered approximately ten months before the implementation of RAS (pre-roll-out: May 2018 – February 2019) and 17 months thereafter (post-roll-out: March 2019 – August 2020).

### Study setting

The study was conducted in three districts in Uganda (Kole, Oyam, Kwania), three Local Government Areas (LGA) in Adamawa State in Nigeria (Fufore, Song, Mayo-Belwa) and three health zones in the Democratic Republic of the Congo (Kenge, Kingandu, Ipamu). The overall study population was 2.5 million of which 476,000 (19%) were children under 5 years. Further details are provided elsewhere [17].

The public health system in the study areas consisted of several levels of community-based providers and at least one level of RHFs (Table 1). Community-based providers implementing pre-referral RAS included CHWs and PHCs. In the study health zones in DRC, CHWs were located in communities with no formal provider within a distance of 5km. In the study LGAs in Nigeria, CHWs were located in communities that were more than 5km away from a public health facility, or the community was hard to reach due to bad road conditions or natural barriers like rivers or mountains. In the study districts in Uganda, there were two CHWs per village irrespective of the presence of other formal health care providers.

**Table 1.**
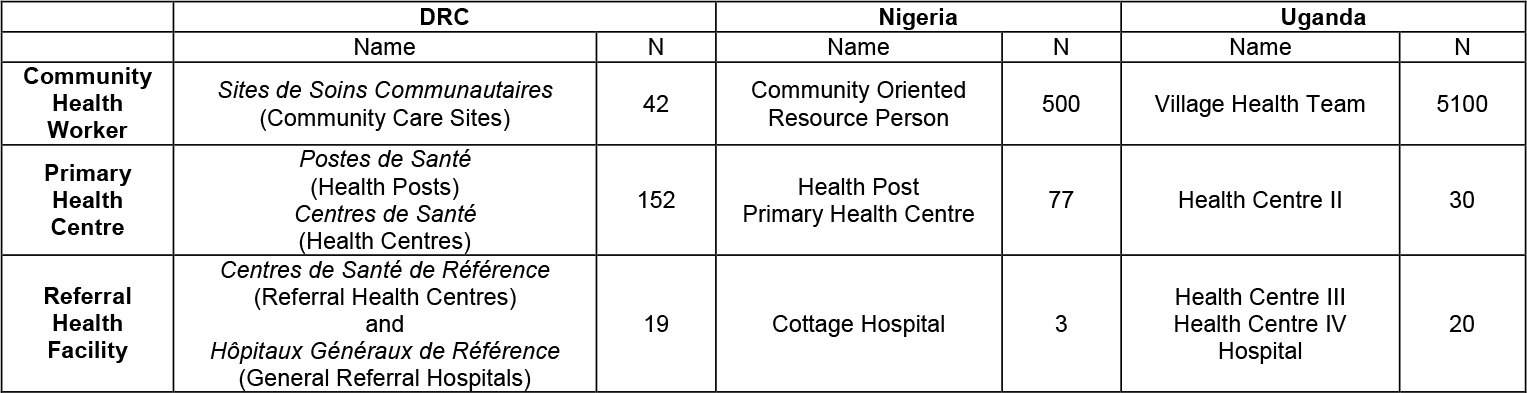
Local names and numbers of community health worker, primary health centres and referral health facilities in 2018, by country.

According to national policies, community-based providers should refer severely ill children to the nearest higher-level health care provider. In the case of CHWs, these are often PHCs (e.g. in Uganda, a Village Health Team may refer a child to a Health Centre II). During RAS roll-out, community-based providers who were trained in the administration of pre-referral RAS were instructed to refer children immediately to a designated RHF. The importance of speedy referral was emphasized in the training. Simultaneously, UNICEF implemented behaviour change communication campaigns that informed caregivers about the benefits of RAS and the importance of referral completion. In Uganda and DRC, there were no interventions in place to support referral to a RHF. In Nigeria, an Emergency Transport System for severely ill children was introduced in July 2019, shortly after the implementation of RAS.

### Data collection

Local research partners established a patient surveillance system for the enrolment and follow-up of children with suspected severe malaria presenting to CHWs or PHCs in DRC and Nigeria, and to CHWs in Uganda. Children were enrolled if they were under five years, had a history of fever and at least one danger sign for which RAS is indicated as per national iCCM and IMCI guidelines. Upon enrolment, community-based providers conducted a malaria rapid diagnostic test (mRDT) for study purposes. After referring the child to a higher-level provider, the enrolling provider reported the case to the local study office where eligible children were recorded in a central case register. In Nigeria and Uganda, this contact happened via telephone. In DRC, community-based providers were regularly visited by CARAMAL research staff to record provisionally enrolled children in the central case register. Dedicated CARAMAL staff were stationed in RHFs in the study area to record the post-referral management of referred children who were admitted for treatment. CARAMAL research staff scheduled an interview 28 days after enrolment. Deceased children were followed up two months after enrolment to respect the mourning period. At the follow-up visit, caregivers or other knowledgeable family members provided information on signs and symptoms, treatment seeking history, diagnosis and treatment during the child’s illness episode. The interviewer also recorded the geo-coordinates of the home location of the child. Additional information on the child’s condition and administered treatment upon enrolment was obtained from the enrolling provider.

Data was collected electronically on tablets with ODK Collect (https://opendatakit.org/). During admission, CARAMAL research staff at RHFs recorded information on case management on paper forms before entering it into ODK Collect. The password protected ODK Aggregate server was hosted at the Swiss Tropical and Public Health Institute in Switzerland.

### Outcomes and explanatory variables

The primary outcome of this analysis was referral completion, defined as a child being brought to one of the designated RHFs at any stage during the treatment seeking process, after seeing a community-based provider, as reported by the caregiver or by CARAMAL staff stationed at the RHF. Secondary outcomes included going to any other public provider after seeing a community-based provider and going to a provider outside of the public health system after seeing a community-based provider. A further secondary outcome was timely referral completion defined as reaching a RHF on the same or next day after enrolment. The number of days between enrolment and reaching a RHF was either calculated as the difference between the enrolment date and the date of admission at a RHF, or obtained from the treatment seeking narrative as reported by the caregiver during follow-up.

The main exposures of interest were the RAS implementation phases (pre-roll-out vs. post-roll-out) and pre-referral RAS administration in the post-roll-out phase. To assess these effects, we grouped children into three study groups: 1) pre-RAS, 2) RAS non-users in the post-RAS phase and 3) RAS users in the post-RAS phase. We accounted for age and sex of the child and the interviewed caregiver, and the child’s place of residence (health zone/LGA/district). The severity as perceived by the caregiver and the presence of a danger sign involving the central nervous system (CNS; convulsions, unusually sleepy, or unconscious) were proxies for disease severity. Additional factors considered included the mRDT result at enrolment, the type of community-based provider (CHW vs. PHC), the season, day of enrolment (workday vs. weekend), enrolment during the Covid-19 pandemic (April 1^st^, 2020 or later), treatment seeking delay between the onset of illness and going to the enrolling provider, the means of transport to enrolling provider, travel time between home and nearest RHF and the administration of home treatment before presentation to enrolling provider.

To calculate the travel time between the home of the child and the nearest RHF, we used the Malaria Atlas Project friction surface 2015 [20] with a 100 m × 100 m resolution. The calculation was done in RStudio [21] using the method described by Bertozzi-Villa [22]. Geolocations of RHFs were obtained from the CARAMAL Health Care Provider Surveys for RHFs within the study area and from Maina et al. [23] for RHFs surrounding the study area. All geolocations were verified using Google Maps [24] and official government sources, where applicable [25, 26].

### Statistical analysis

For each country, we used a logistic regression model to estimate the association of the implementation of RAS and RAS administration with referral completion. For children completing referral, we used logistic regression models to estimate the association between RAS implementation and administration and referral timeliness. All models included the enrolling PHC or CHW as random effects to account for clustering at that level. Exposure variables were selected based on rational grounds prior to analysis and included in the final model irrespective of their level of significance. Variables to test for interactions were chosen *a priori*. The interactions included in the final model were significant at the 5% level. Observations with missing values for referral completion were excluded from the regression analysis. Statistical analyses were performed in Stata SE 16.1 [27].

### Ethics

The community-based providers informed caregivers about the CARAMAL study prior to enrolment and caregivers gave oral pre-consent to be contacted for a follow-up interview. We obtained written consent from all caregivers of provisionally enrolled children either at the RHF or before the follow-up interview 28 days after enrolment.

The CARAMAL study protocol was approved by the Research Ethics Review Committee of the World Health Organization (WHO ERC, No. ERC.0003008), the Ethics Committee of the University of Kinshasa School of Public Health (No. 012/2018), the Health Research Ethics Committee of the Adamawa State Ministry of Health (S/MoH/1131/I), the National Health Research Ethics Committee of Nigeria (NHREC/01/01/2007-05/05/2018), the Higher Degrees, Research and Ethics Committee of the Makerere University School of Public Health (No. 548), the Uganda National Council for Science and Technology (UNCST, No. SS 4534), and the Scientific and Ethical Review Committee of CHAI (No. 112, 21 Nov 2017). The study is registered on ClinicalTrials.gov (NCT03568344).

## Results

### Study population

Between May 2018 and August 2020, community-based providers provisionally enrolled 8,365 children (Supplement 1). The study team successfully followed up 7,593 (91%) children and obtained informed consent. Of those, 6,505 (78%) children fulfilled all inclusion criteria of which 6,449 had a known referral status (77%). The majority of included children were enrolled after the implementation of RAS (n=4,396, 68%). Particularly in DRC, the sample receiving RAS in the post-RAS phase was substantially larger (N = 1,548) compared to the other two study groups (pre-RAS: N = 368; post-RAS non-users: N = 188) (Table 2). In Nigeria, the sample sizes across study groups were balanced. In Uganda, the number of children enrolled in the pre-RAS phase (N = 1,479) was comparable to the number of children receiving RAS in the post-RAS phase (N = 1,631); however, the number of children not receiving RAS in the post-RAS phase was substantially smaller (N = 635).

**Table 2.**
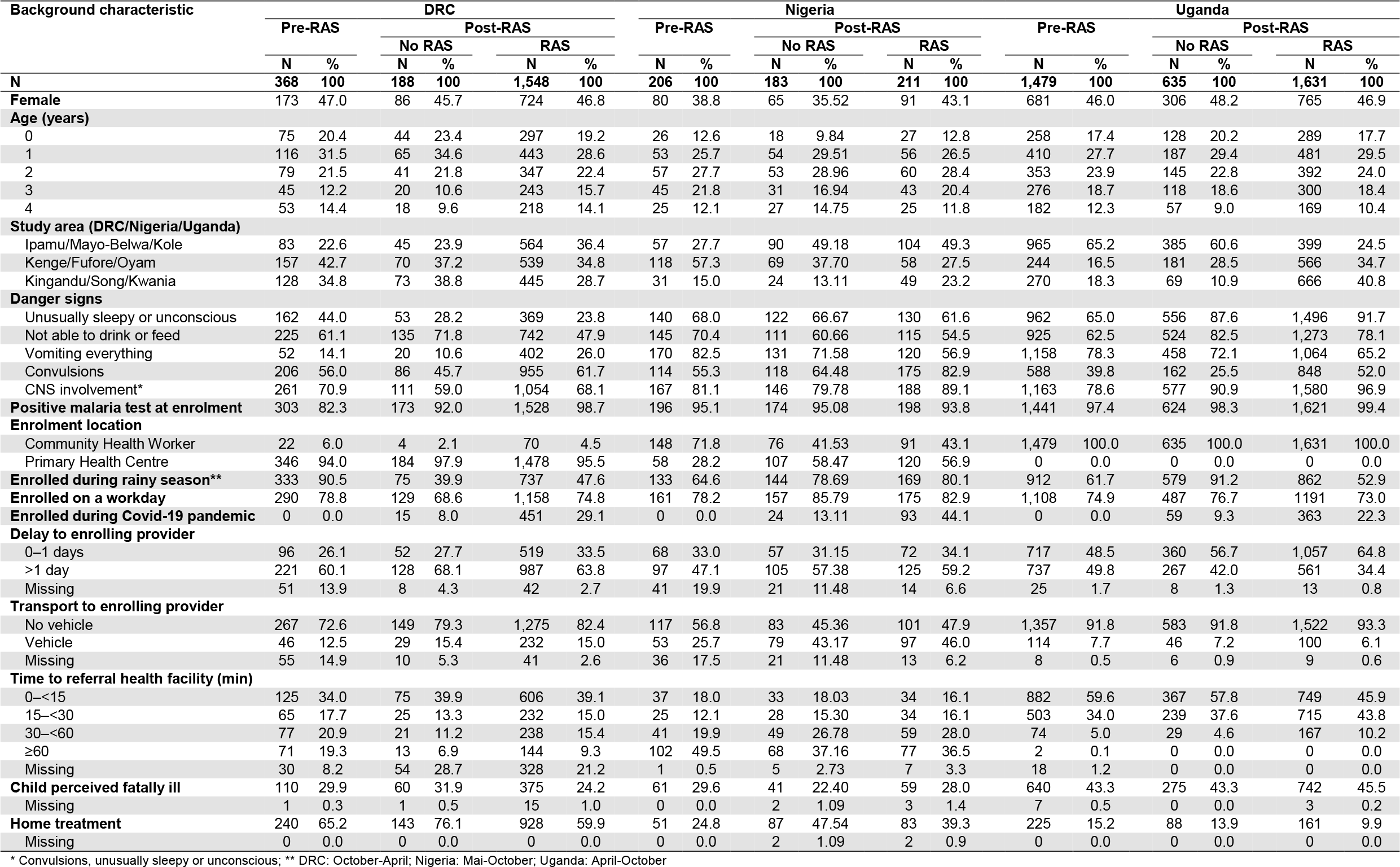
Study population characteristics by country and study group.

Within each country, the age and sex distribution were similar among children enrolled in the pre-RAS phase, RAS users and non-users in the post-RAS phase. In Nigeria, there were fewer children under one year than in the other countries. Danger signs involving the CNS were most common in Uganda followed by Nigeria and DRC. In all countries, more than 90% of eligible children tested positive for malaria at enrolment. In DRC, eligible children were almost exclusively enrolled by PHCs (95%), while in Uganda all children were enrolled by CHWs. In Nigeria, a higher proportion of children were enrolled by CHWs in the pre-RAS phase (72%) compared to the post-RAS phase (42%). In the post-RAS phase, between 19% (Uganda) and 30% (Nigeria) of the children were enrolled during the Covid-19 pandemic. In all countries, the proportion of children receiving RAS was higher during the Covid-19 pandemic.

### Referral completion

In DRC, overall 1,408 (67%) children completed referral to a designated RHF. Few children went to another public provider (6%) or to any other provider (3%). In Nigeria, 287 (48%) children completed referral to a designated RHF. An additional 21% of the children went to another public provider and only 4% of the children went to a non-public provider. In Uganda, 2,170 (58%) of the children completed referral to the designated RHFs. Going to another public provider was infrequent (9%), but 29% of the children in Uganda were brought to providers outside of the public health system.

In all countries, referral completion to a RHF was slightly lower among RAS users compared to RAS non-users in the post-RAS phase (Figure 1, Table 3 and Table 4). In DRC and Uganda, referral completion in the post-RAS phase was comparable to referral completion in the pre-RAS phase. Meanwhile in Nigeria, referral completion increased from the pre-RAS to the post-RAS phase. The difference between the pre-RAS and post-RAS phase was mainly driven by PHC enrolments being substantially more likely to complete referral to a RHF than CHW enrolments, in combination with an increase in the number of PHC enrolments in the post-RAS phase (Figure 2).

**Figure 1.**
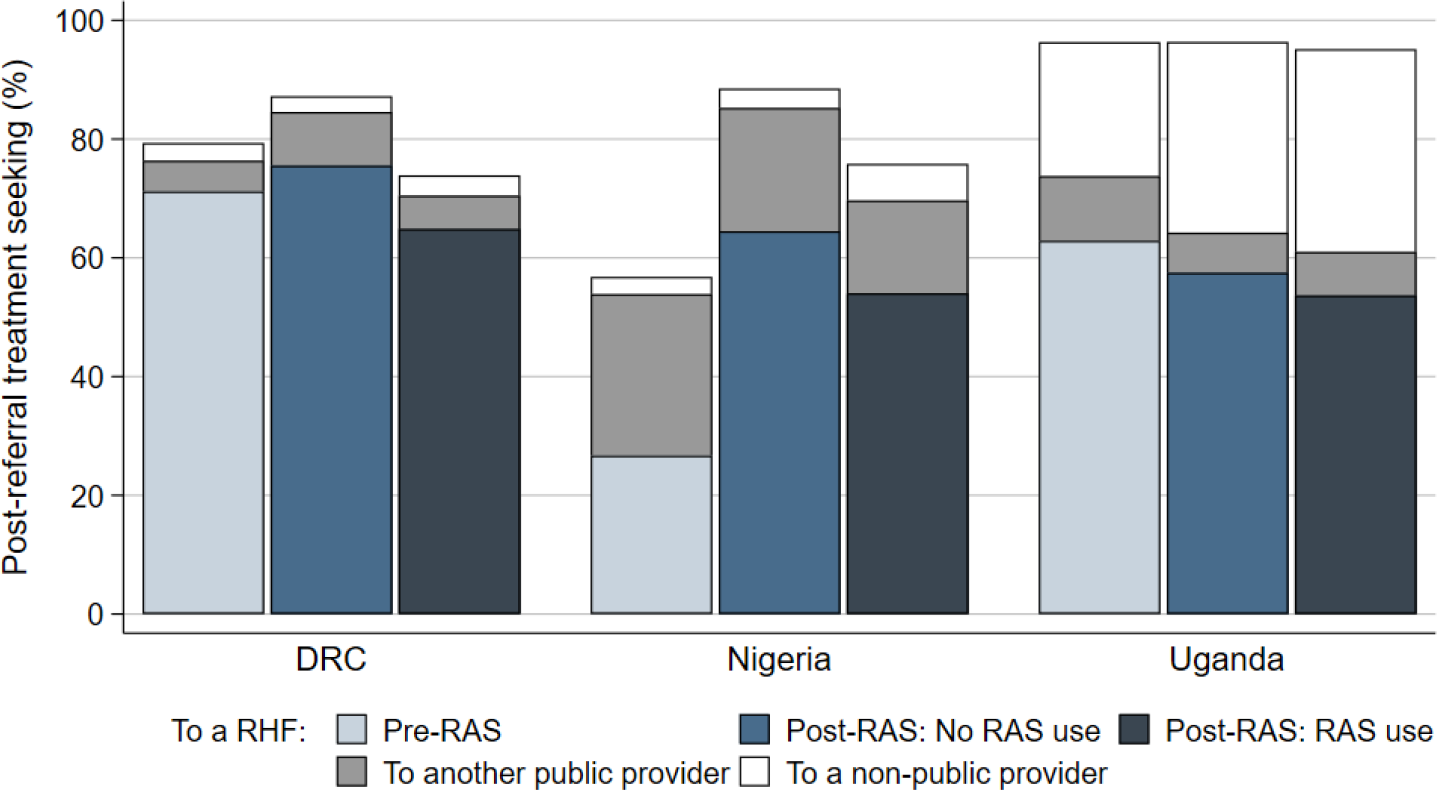
Post-referral treatment seeking by country and study group.

**Table 3.** Estimated associations between selected factors and referral completion in DRC and Uganda.

|  | DRC |  |  |  |  |  | Uganda |  |  |  |  |  |
| --- | --- | --- | --- | --- | --- | --- | --- | --- | --- | --- | --- | --- |
|  | n | N | % | Adjusted OR§ | 95% CI | p-value | n | N | % | Adjusted OR§ | 95% CI | p-value |
| **All** | **1,408** | **2,104** | **66.9** |  |  |  | **2,170** | **3,745** | **57.9** |  |  |  |
| **Study group** |  |  |  |  |  |  |  |  |  |  |  |  |
| Pre-RAS | 262 | 368 | 71.2 | Ref. |  |  | 930 | 1,479 | 62.9 | Ref. |  |  |
| Post-RAS |  |  |  |  |  |  |  |  |  |  |  |  |
| No RAS use | 142 | 188 | 75.5 | 0.34 | 0.18–0.66 | 0.001 | 365 | 635 | 57.5 | 0.80 | 0.63–1.01 | 0.06 |
| RAS use | 1,004 | 1,548 | 64.9 | 0.48 | 0.30–0.77 | 0.002 | 875 | 1,631 | 53.6 | 0.72 | 0.58–0.88 | 0.002 |
| **Enrolment location** |  |  |  |  |  |  |  |  |  |  |  |  |
| CHW | 42 | 96 | 43.8 | Ref. |  |  | 2,170 | 3,745 | 57.9 |  |  |  |
| PHC | 1,366 | 2,008 | 68.0 | 4.85 | 1.22–19.25 | 0.02 | 0 | 0 | NA | NA |  |  |
| **CNS danger sign\*** |  |  |  |  |  |  |  |  |  |  |  |  |
| No | 532 | 678 | 78.5 | Ref. |  |  | 267 | 425 | 62.8 | Ref. |  |  |
| Yes | 876 | 1,426 | 61.4 | 0.58 | 0.41–0.82 | 0.002 | 1,903 | 3,320 | 57.3 | 0.80 | 0.61–1.04 | 0.09 |
| **Enrolled during rainy season\*\*** |  |  |  |  |  |  |  |  |  |  |  |  |
| No | 665 | 959 | 69.3 | Ref. |  |  | 772 | 1,392 | 55.5 | Ref. |  |  |
| Yes | 743 | 1,145 | 64.9 | 0.77 | 0.56–1.07 | 0.12 | 1,398 | 2,353 | 59.4 | 1.15 | 0.97–1.38 | 0.11 |
| **Enrolled on a workday** |  |  |  |  |  |  |  |  |  |  |  |  |
| No | 365 | 527 | 69.3 | Ref. |  |  | 516 | 959 | 53.8 | Ref. |  |  |
| Yes | 1,043 | 1,577 | 66.1 | 0.91 | 0.65–1.29 | 0.60 | 1,654 | 2,786 | 59.4 | 1.19 | 1.00–1.41 | 0.05 |
| **Enrolled during Covid-19 pandemic** |  |  |  |  |  |  |  |  |  |  |  |  |
| No | 1,092 | 1,638 | 66.7 | Ref. |  |  | 1,922 | 3,323 | 57.8 | Ref. |  |  |
| Yes | 316 | 466 | 67.8 | 1.15 | 0.78–1.70 | 0.48 | 248 | 422 | 58.8 | 0.90 | 0.69–1.19 | 0.48 |
| **Delay to enrolling provider** |  |  |  |  |  |  |  |  |  |  |  |  |
| 0–1 days | 402 | 667 | 60.3 | Ref. |  |  | 1,212 | 2,134 | 56.8 | Ref. |  |  |
| > 1 day | 915 | 1,336 | 68.5 | 1.07 | 0.77–1.49 | 0.69 | 925 | 1,565 | 59.1 | 1.14 | 0.97–1.34 | 0.12 |
| Missing | 91 | 101 | 90.1 | 3.33 | 0.86–12.83 | 0.08 | 33 | 46 | 71.7 | 1.11 | 0.43–2.86 | 0.83 |
| **Transport to enrolling provider** |  |  |  |  |  |  |  |  |  |  |  |  |
| No vehicle | 1,065 | 1,691 | 63.0 | Ref. |  |  | 2,002 | 3,462 | 57.8 | Ref. |  |  |
| Vehicle | 251 | 307 | 81.8 | 1.08 | 0.66–1.79 | 0.75 | 150 | 260 | 57.7 | 1.06 | 0.78–1.42 | 0.72 |
| Missing | 92 | 106 | 86.8 | 4.80 | 1.37–16.84 | 0.01 | 18 | 23 | 78.3 | 3.31 | 0.78–14.06 | 0.11 |
| **Time to referral health facility (min)** |  |  |  |  |  |  |  |  |  |  |  |  |
| 0–<15 | 668 | 806 | 82.9 | Ref. |  |  | 1,349 | 1,998 | 67.5 | Ref. |  |  |
| 15–<30 | 232 | 322 | 72.0 | 1.12 | 0.65–1.95 | 0.68 | 692 | 1,457 | 47.5 | 0.72 | 0.58–0.89 | 0.003 |
| 30–<60 | 228 | 336 | 67.9 | 0.80 | 0.46–1.40 | 0.44 | 115 | 270 | 42.6 | 0.55 | 0.39–0.79 | 0.001 |
| ≥60 | 80 | 228 | 35.1 | 0.46 | 0.24–0.89 | 0.02 | 0 | 2 | 0.0 | NA |  |  |
| Missing | 200 | 412 | 48.5 | 0.87 | 0.52–1.46 | 0.60 | 14 | 18 | 77.8 | 1.88 | 0.53–6.67 | 0.33 |
| **Perceived severity** |  |  |  |  |  |  |  |  |  |  |  |  |
| Not fatal | 979 | 1,542 | 63.5 | Ref. |  |  | 1,180 | 2,078 | 56.8 | Ref. |  |  |
| Fatal | 413 | 545 | 75.8 | 1.86 | 1.28–2.71 | 0.001 | 985 | 1,657 | 59.4 | 1.11 | 0.94–1.30 | 0.21 |
| Missing | 16 | 17 | 94.1 | 16.14 | 0.61–429.60 | 0.10 | 5 | 10 | 50.0 | 0.80 | 0.20–3.19 | 0.75 |
| **Home treatment** |  |  |  |  |  |  |  |  |  |  |  |  |
| No | 445 | 793 | 56.1 | Ref. |  |  | 1,876 | 3,271 | 57.4 | Ref. |  |  |
| Yes | 963 | 1,311 | 73.5 | 1.43 | 1.03–1.99 | 0.03 | 294 | 474 | 62.0 | 1.10 | 0.87–1.39 | 0.41 |
| Missing | 0 | 0 | NA | NA |  |  | 0 | 0 | NA | NA |  |  |

\* Danger signs involving the central nervous system (CNS): convulsions, unusually sleepy or unconscious
\*\* DRC: October–April; Uganda: April–October
§ Odds ratio additionally adjusted for child sex, child age, caregiver sex, caregiver age, location of residence (health zone, district), and malaria test result.

**Table 4.** Estimated associations between selected factors and referral completion in Nigeria.

|  | Nigeria |  |  |  |  |  |
| --- | --- | --- | --- | --- | --- | --- |
|  | n | N | % | Adjusted OR§ | 95% CI | p-value |
| **All** | **287** | **600** | **47.8** |  |  |  |
| **Study group by enrolment location** |  |  |  |  |  |  |
| **CHW** |  |  |  |  |  |  |
| Pre-RAS | 9 | 148 | 6.1 | Ref. |  |  |
| Post-RAS |  |  |  |  |  |  |
| No RAS use | 16 | 76 | 21.1 | 3.97 | 1.07–14.75 | 0.04 |
| RAS use | 24 | 91 | 26.4 | 9.95 | 2.71–36.58 | <0.001 |
| **PHC\*** |  |  |  |  |  |  |
| Pre-RAS | 46 | 58 | 79.3 | Ref. |  |  |
| Post-RAS |  |  |  |  |  |  |
| No RAS use | 102 | 107 | 95.3 | 35.09 | 6.52–188.75 | <0.001 |
| RAS use | 90 | 120 | 75.0 | 6.45 | 1.62–25.67 | 0.01 |
| **Enrolment location** |  |  |  |  |  |  |
| CHW | 49 | 315 | 15.6 | Ref. |  |  |
| PHC* | 238 | 285 | 83.5 | 19.79 | 2.97–131.71 | 0.002 |
| **CNS involvement\*\*** |  |  |  |  |  |  |
| No | 33 | 99 | 33.3 | Ref. |  |  |
| Yes | 254 | 501 | 50.7 | 0.48 | 0.17–1.38 | 0.17 |
| **Enrolled during rainy season\*\*\*** |  |  |  |  |  |  |
| No | 78 | 154 | 50.6 | Ref. |  |  |
| Yes | 209 | 446 | 46.9 | 0.70 | 0.29–1.67 | 0.42 |
| **Enrolled on a workday** |  |  |  |  |  |  |
| No | 33 | 107 | 30.8 | Ref. |  |  |
| Yes | 254 | 493 | 51.5 | 0.74 | 0.31–1.76 | 0.50 |
| **Enrolled during Covid-19 pandemic** |  |  |  |  |  |  |
| No | 239 | 483 | 49.5 | Ref. |  |  |
| Yes | 48 | 117 | 41.0 | 0.09 | 0.03–0.26 | <0.001 |
| **Delay to enrolling provider** |  |  |  |  |  |  |
| 0–1 days | 90 | 197 | 45.7 | Ref. |  |  |
| > 1 day | 154 | 327 | 47.1 | 1.64 | 0.77–3.52 | 0.20 |
| Missing | 43 | 76 | 56.6 | 3.53 | 1.03–12.09 | 0.04 |
| **Transport to enrolling provider** |  |  |  |  |  |  |
| No vehicle / missing**** | 125 | 371 | 33.7 | Ref. |  |  |
| Vehicle | 162 | 229 | 70.7 | 0.82 | 0.32–2.10 | 0.68 |
| **Time to referral health facility (min)** |  |  |  |  |  |  |
| 0–<15 | 85 | 104 | 81.7 | Ref. |  |  |
| 15–<30 | 62 | 87 | 71.3 | 0.48 | 0.13–1.73 | 0.26 |
| 30–<60 | 83 | 149 | 55.7 | 0.23 | 0.07–0.77 | 0.02 |
| ≥60 | 48 | 247 | 19.4 | 0.06 | 0.02–0.22 | <0.001 |
| Missing | 9 | 13 | 69.2 | 0.13 | 0.01–1.84 | 0.13 |
| **Perceived severity** |  |  |  |  |  |  |
| Not fatal | 212 | 434 | 48.8 | Ref. |  |  |
| Fatal | 72 | 161 | 44.7 | 0.63 | 0.30–1.32 | 0.23 |
| Missing | 3 | 5 | 60.0 | 1.23 | 0.01–153.00 | 0.93 |
| **Home treatment** |  |  |  |  |  |  |
| No / missing**** | 166 | 379 | 43.8 | Ref. |  |  |
| Yes | 121 | 221 | 54.8 | 1.08 | 0.54–2.16 | 0.82 |

\* Adjusted for LGA. Odds ratio shown for Mayo-Belwa. Odds ratios for Fufore and Song are higher.
\*\* Danger signs involving the central nervous system (CNS): convulsions, unusually sleepy or unconscious
\*\*\* Mai-October
\*\*\*\* Observations with missing values added to reference category because no meaningful odds ratio could be computed due to the data structure (missing values in other covariates).
§ Odds ratio additionally adjusted for child sex, child age, caregiver sex, caregiver age, location of residence (LGA), and malaria test result.

**Figure 2.**
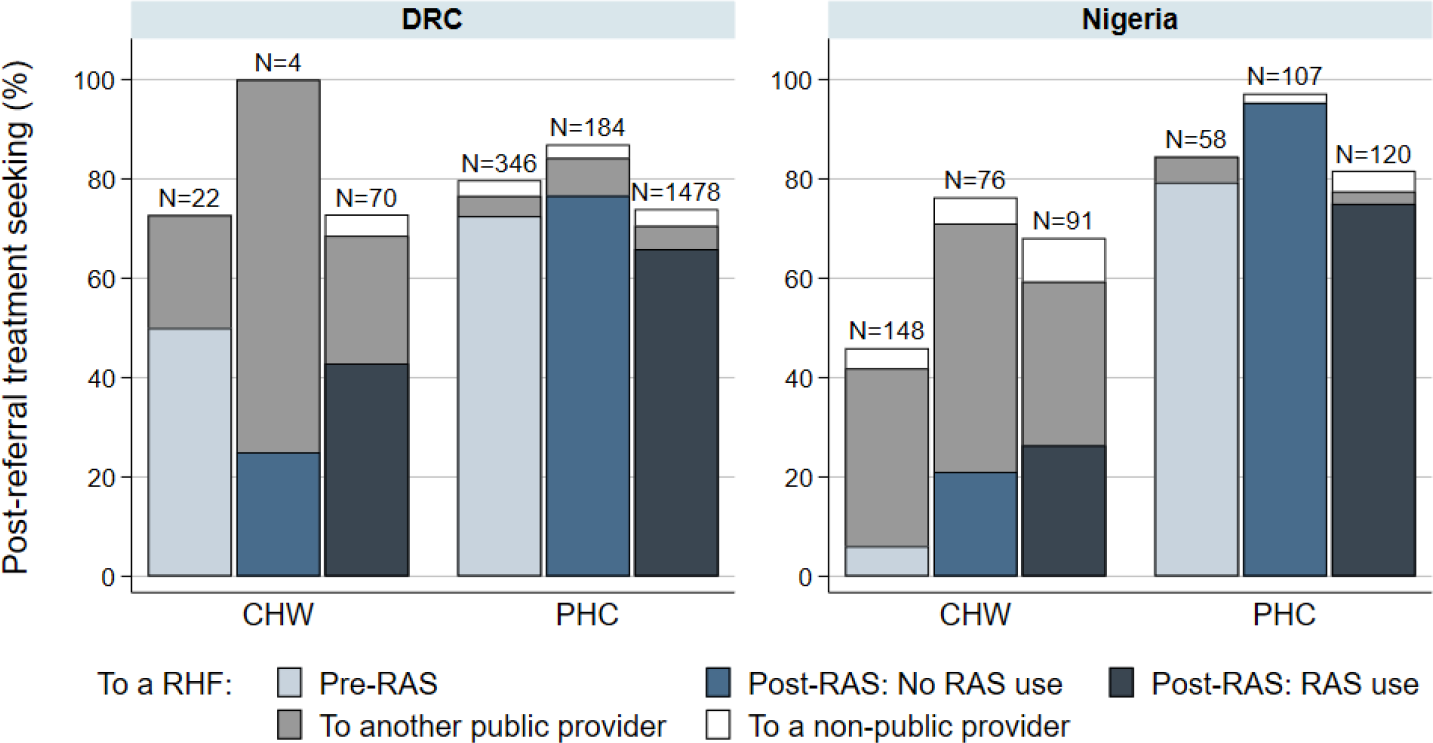
Post-referral treatment seeking by enrolment location and study group, in DRC and Nigeria.

In both DRC and Nigeria, going to any other provider than a RHF was uncommon for PHC enrolments (Figure 2); but seemed more common for CHW enrolments who frequently went to a public provider other than a designated RHF. In Uganda, children rarely went to another public provider but tended instead to go to a private provider, compensating the lower referral completion to a RHF in the two post-RAS study groups (Figure 1).

In DRC and Uganda, referral completion was lower in the post-RAS phase compared to the pre-RAS phase after adjusting for other factors, irrespective of whether children had received RAS (Table 3). The opposite occurred in Nigeria, where referral completion in the post-RAS phase irrespective of RAS use was higher compared to the pre-RAS phase (Table 4).

When taking RAS non-users in the post-RAS phase as a reference, the odds of completing referral did not significantly differ between children not receiving RAS and children receiving RAS in DRC (adjusted odds ratio (aOR) = 1.39, 95% confidence interval (CI) 0.82–2.35) and Uganda (aOR = 0.90, 95% CI 0.70–1.16). In Nigeria, the same was true for children enrolled by a CHW (aOR = 2.51, 95% CI 0.76–8.24); however, among children enrolled by a PHC, those who had received RAS were significantly less likely to complete referral than those not receiving RAS in the post-RAS phase (aOR = 0.18, 95% CI 0.05–0.71).

Besides RAS implementation and RAS administration, we found other factors significantly associated with referral completion. In all countries, increasing travel time to the RHF had a negative effect on referral completion. In DRC and Nigeria, children were more likely to complete referral if they were referred by a PHC compared to a CHW (not applicable in Uganda). Other factors that had a positive effect on referral completion included being perceived fatally ill by the caregiver (DRC), having received home treatment (DRC) or being enrolled on a workday (Uganda). Factors with a negative effect on referral completion included having a CNS danger sign (DRC) or being enrolled during the Covid-19 pandemic (Nigeria).

The adjusted odds ratios did not differ substantially from the unadjusted estimates except for the effect of the malaria test result in DRC and being enrolled by a PHC in Nigeria. Unadjusted estimates are provided in Supplement 2.

### Referral timeliness

Of the 3,865 children that completed referral to a RHF, data on the timeliness of referral completion was available for 3,598 children (93%) (Supplement 1). Timely referral completion to a RHF on the same or next day after seeing a community-based provider was 76% in DRC, 86% in Nigeria and 92% in Uganda. In all countries, timely referral was highest among children receiving RAS in the post-RAS phase; however, the differences between study groups were rather small (Figure 3). After adjusting for other factors, children in Uganda receiving RAS in the post-RAS phase were significantly more likely to complete referral on time than children in the pre-RAS phase (OR = 1.81, 95% CI 1.17–2.79). Other comparisons between study groups were not significant in any of the countries. Complete tables with denominators and regression results are presented in Supplement 3.

**Figure 3.**
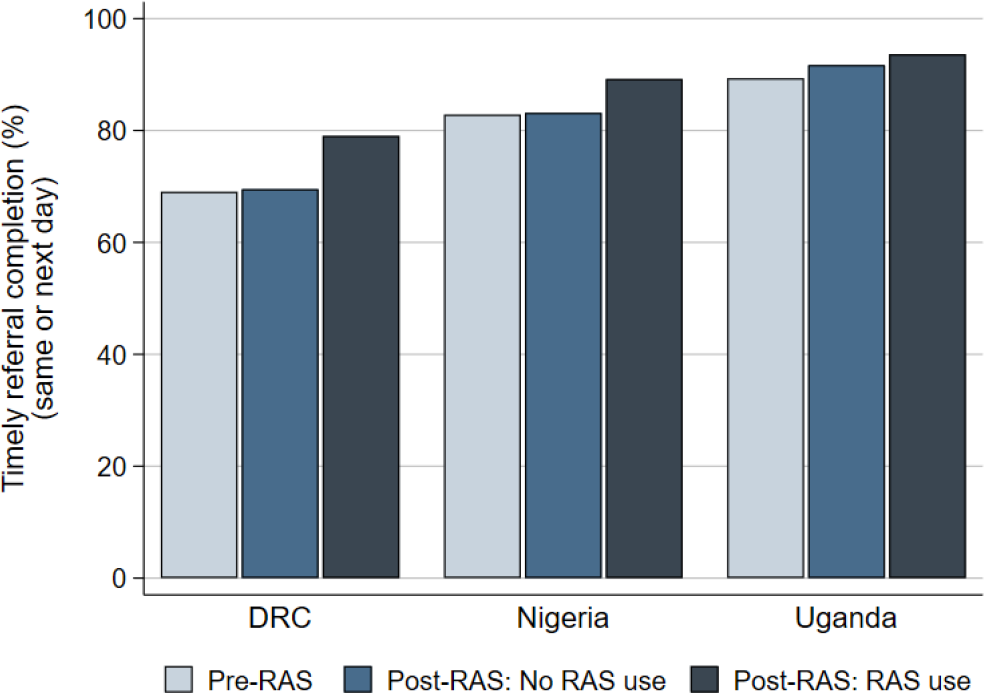
Timely referral completion, by country and study group, on the same or next day after referral by a community-based provider, from all patients completing referral to a referral health facility.

## Discussion

After the administration of pre-referral RAS, current guidelines recommend referral completion to a health facility where intramuscular or intravenous treatment is available [5]. Findings from previous studies on RAS and referral completion were mostly reassuring; however, the effect of RAS on referral completion was either not adjusted for other factors [6] or did not compare RAS users to non-users [8-11, 13]. Additionally, none of the studies took into consideration that the nearest health facility might not have the capacity of administering parenteral antimalarial treatment. The CARAMAL Project for the first time provides adjusted estimates of the effect of RAS on referral completion to a RHF at which, according to national policy, appropriate post-referral treatment is available. Post-referral treatment with an injectable antimalarial followed by a full course of ACT ensures that children are effectively treated for severe malaria, and RAS (and parenteral artemisinin) is not applied as a monotherapy, thereby reducing the risk of the development and selection of artemisinin-resistant parasites [5].

This study provides evidence that the large-scale introduction and use of RAS can affect referral completion. In DRC and Uganda, referral completion was lower in the post-roll-out phase compared to the pre-roll-out phase. In Nigeria, the opposite was the case. However, in Nigeria, children who were administered RAS in a PHC were less likely to complete referral to a RHF than children who did not receive RAS. Referral completion by children attending a PHC in Nigeria and DRC was consistently higher when compared to the referral completion of children attending a CHW (not applicable in Uganda). In all countries, children living a greater distance from a RHF (measured in the time it would take to travel to the facility) were significantly less likely to complete referral than those living in the vicinity of the facility. In all countries, the majority of children that completed referral to a RHF did so on the same or next day after being referred.

The findings of this study raise legitimate concerns that the roll-out of RAS may lead to lower referral completion in children who were administered pre-referral RAS. The comparison of RAS users and RAS non-users in the post-roll-outs phase in Nigeria strongly suggests that RAS administration negatively affected referral completion of children enrolled in a PHC. In DRC, a comparison of RAS users versus non-users in the post-roll-out phase was limited by the comparatively small number of non-users. Therefore, the significantly lower referral completion of RAS users compared to the referral completion of children enrolled in the pre-roll-out phase may indicate that referral completion was negatively affected by RAS administration; even though other causal links cannot be excluded. The reason for a decrease in referral completion after RAS use is most likely the rapid improvement of children after RAS administration, a result of the fast reduction of the parasite to blood concentration and the drug’s antipyretic effect [28-31]. Considering that treatment seeking is often delayed due to lack of transport and money [32-34], the child’s condition may have improved in the meantime. In such a situation, a caregiver is likely to balance between the referral recommendation and priorities at home, in addition to expenses for transport and those that would be incurred at the RHF [4]. Obviously, such a practice raises the concern that children may not receive the appropriate post-referral treatment with potentially fatal consequences.

Measures to improve referral completion must take into consideration the actual capacity of RHFs to provide appropriate case management for severe malaria. As opposed to previous studies, this study considered referral to be completed only if the patient arrived at a RHF with the capacity to manage severe malaria cases. However, an analyses of the quality of care at RHFs in the CARAMAL study areas found that the treatment of children with severe malaria was often inadequate [19]. Meanwhile, some children sought post-referral treatment from a lower-level public or from a non-public provider. Considering that referral completion did not improve the health outcome of children enrolled in the CARAMAL study in DRC and Uganda [18] sufficient treatment may have been provided by non-RHF providers [35]. It is also possible that less severely sick children recover faster and are hence less likely to be brought to a RHF after pre-referral RAS treatment. Therefore, children with suspected severe malaria first attending a community-based provider may not always require treatment at the level of RHFs. However, recognizing such children with a more moderate form of severe malaria remains a challenge.

The finding that a substantial proportion of children (33–52%) did not complete referral emphasizes the need to address referral-related barriers at all levels: sensitizing caregivers, properly training community-based health care providers, facilitating access to and increasing trust in RHFs. Such a package of supportive interventions accompanying the roll-out of RAS in Zambia has previously been shown to reduce the mortality of children with severe malaria [36]. In a trial conducted by Gomes et al. [1], RAS had a protective effect in the context of high completion of referral (though not necessarily to a RHF in the African sites). In the CARAMAL study in Nigeria, the implementation of an Emergency Transport System most likely increased referral completion, and referral completion significantly improved the health outcomes of children with suspected severe malaria in this country. The available evidence strongly indicates that community-based programmes should always be accompanied by measures strengthening referral.

Irrespective of the effort to strengthen referral, it is important to acknowledge that some caregivers to children may delay or not complete referral. Therefore, the training materials and referral guidelines for community-based providers need to emphasize the importance of a close follow-up of severely sick children, if necessary at their home. If referral cannot be completed or is refused by the caregiver, the treatment with pre-referral drugs should be continued. Such a recommendation already exists for RAS until oral treatment with an ACT is tolerated [2]. Similarly, the WHO recommends that CHWs continue administering amoxicillin to children with pneumonia with chest in-drawing if referral is not feasible [37]. Sufficient stocks of pre-referral drugs are therefore essential for providing adequate care to children unable to complete referral.

Referral completion to a RHF was a problem particularly among CHW enrolments in DRC and Nigeria. Unlike in Uganda, CHWs in these two countries are placed in especially hard-to-reach areas. In our analyses, we accounted for difficulties in geographical access to a RHF (availability of transport and travel time to RHF), but we may have missed additional barriers to accessing a RHF. Another explanation could be that children attending a PHC are more severely ill than children attending a CHW [38]. Caregivers may be more likely to make increased efforts to reach the first provider as well as to complete referral if the child is more severely ill. Meanwhile, irrespective of the reasons, an active follow up of children at home seems to be particularly important in the most hard-to-reach places where referral completion is the least likely. As community programmes continue to be the preferred approach to extend health services to remote communities, the challenges associated with these hard-to-reach places need to be acknowledged in referral and treatment guidelines and the promotion of best practices.

Even though treatment seeking practices including referral are highly contextual, some recommendations based on the results of this study can be generalised to other settings. That is, programmes implementing RAS need to consider the potential effects on referral completion. More generally, community-based programmes should be supported by measures facilitating referral completion, and provide a back-up option for those children who fail to complete referral. Alternative treatment options are particularly important in hard-to-reach places.

This study has several strengths. First, it covered three different contexts with varying intensities of malaria transmission, access to health care, and differences in the implementation of iCCM/IMCI policies [17]. Secondly, the study was community-based and enrolled a large number of children with severe febrile illness from remote communities. Large community-based studies in far-to-reach places are mostly cross-sectional surveys that rarely capture severe illness episodes because of their low incidence, and always exclude children that are deceased, resulting in a lack of understanding of severe illnesses at community level [39]. Thirdly, the study achieved a high follow-up rate, thereby reducing the risk of selection bias.

This study comes with several limitations. First, the low enrolment numbers of children not receiving RAS in the post-roll-out phase did not allow a clear conclusion about the effects of RAS administration on referral completion after the roll-out of RAS in DRC. Secondly, the enrolment strategy in Nigeria and Uganda may have introduced selection bias. The notification of enrolments from the enrolling provider to the local study office depended on a contact via mobile phone. Thus, the study may have excluded systematically children in the most remote places because of unstable network coverage. It is likely that these children would have also been the least likely to complete referral leading to an overestimation of referral completion in our study. Thirdly, the observational design and the retrospective data collection 28 days after enrolment did not allow for direct causal inferences.

## Conclusion

Providing prompt and appropriate health care to severely sick children in remote communities remains a challenge. Children in hard-to-reach places are the least likely to complete referral after seeing a community-based provider. In addition, referral completion may further be negatively affected by the administration of RAS. To ensure that community-based programmes are effectively implemented, barriers to referral completion need to be addressed at all levels. Alternative effective treatment options should be provided to children unable to complete referral.

## Supporting information

STROBE checklist

## Data Availability

Individual de-identified participant data that underlie the results reported in this article are available at zenodo.org upon reasonable request (DOI: 10.5281/zenodo.5570278).

## Acknowledgements

The authors would like to express their sincere thanks to the children and their caregivers who agreed to participate in this study; the health workers and local and national health authorities who provided their support; the study teams of the School of Public Health in Kinshasa (DRC), Akena Associates (Nigeria), and Makerere University School of Public Health (Uganda); and the colleagues of the local CHAI and UNICEF offices. We greatly appreciate Aurelio DiPasquale’s support with the ODK software and Robert Canavan’s editorial support.

## Supporting information captions

**Supplement 1**. Inclusion flow-charts

**Supplement 2**. Table S2.1. Estimated associations between child characteristics and referral completion, DRC.

**Supplement 2**. Table S2.2. Estimated associations between child characteristics and referral completion, Nigeria.

**Supplement 2**. Table S2.3. Estimated associations between child characteristics and referral completion, Uganda.

**Supplement 3**. Table S3 1. Estimated associations between child characteristics and referral timeliness, DRC.

**Supplement 3**. Table S3 2. Estimated associations between child characteristics and referral timeliness, Nigeria.

**Supplement 3**. Table S3 3. Estimated associations between child characteristics and referral timeliness, Uganda.

## References

1. Gomes MF, Faiz MA, Gyapong JO, Warsame M, Agbenyega T, Babiker A, et al. Prereferral rectal artesunate to prevent death and disability in severe malaria: a placebo-controlled trial. Lancet. 2009;373(9663):557–66. doi: 10.1016/S0140-6736(08)61734-1.

2. World Health Organization. Rectal artesunate for pre-referral treatment of severe malaria. 2017.

3. Gomes M, Ribeiro I, Warsame M, Karunajeewa H, Petzold M. Rectal artemisinins for malaria: a review of efficacy and safety from individual patient data in clinical studies. BMC Infect Dis. 2008;8:39. Epub 2008/04/01. doi: 10.1186/1471-2334-8-39. PubMed PMID: 18373841; PubMed Central PMCID: PMCPMC2364627.

4. Simba DO, Kakoko DC, Warsame M, Premji Z, Gomes MF, Tomson G, et al. Understanding caretakers’ dilemma in deciding whether or not to adhere with referral advice after pre-referral treatment with rectal artesunate. Malar J. 2010;9:123. doi: 10.1186/1475-2875-9-123.

5. World Health Organization. Guidelines for the treatment of malaria - 3rd edition. Geneva: World Health Organization, 2015.

6. Mvumbi PM, Musau J, Faye O, Situakibanza H, Okitolonda E. Adherence to the referral advice after introduction of rectal artesunate for pre-referral treatment of severe malaria at the community level: a noninferiority trial in the Democratic Republic of the Congo. Malar J. 2019;18(1):438. doi: 10.1186/s12936-019-3074-6.

7. Strachan CE, Nuwa A, Muhangi D, Okui AP, Helinski MEH, Tibenderana JK. Community understanding of the concept of pre-referral treatment and how this impacts on referral related decision-making following the provision of rectal artesunate: a qualitative study in western Uganda. BMC Health Serv Res. 2018;18(1):470. doi: 10.1186/s12913-018-3209-4.

8. Simba DO, Warsame M, Kimbute O, Kakoko D, Petzold M, Tomson G, et al. Factors influencing adherence to referral advice following pre-referral treatment with artesunate suppositories in children in rural Tanzania. Trop Med Int Health. 2009;14(7):775–83. doi: 10.1111/j.1365-3156.2009.02299.x.

9. Siribié M, Ajayi IO, Nsungwa-Sabiiti J, Sanou AK, Jegede AS, Afonne C, et al. Compliance With Referral Advice After Treatment With Prereferral Rectal Artesunate: A Study in 3 Sub-Saharan African Countries. Clin Infect Dis. 2016;63(Suppl 5):S283–S9. doi: 10.1093/cid/ciw627.

10. Ajayi IO, Nsungwa-Sabiiti J, Siribié M, Falade CO, Sermé L, Balyeku A, et al. Feasibility of Malaria Diagnosis and Management in Burkina Faso, Nigeria, and Uganda: A Community-Based Observational Study. Clin Infect Dis. 2016;63(Suppl 5):S245–S55. doi: 10.1093/cid/ciw622.

11. Lal S, Ndyomugenyi R, Paintain L, Alexander ND, Hansen KS, Magnussen P, et al. Caregivers’ compliance with referral advice: evidence from two studies introducing mRDTs into community case management of malaria in Uganda. BMC Health Serv Res. 2018;18(1):317. doi: 10.1186/s12913-018-3124-8.

12. Nanyonjo A, Bagorogoza B, Kasteng F, Ayebale G, Makumbi F, Tomson G, et al. Estimating the cost of referral and willingness to pay for referral to higher-level health facilities: a case series study from an integrated community case management programme in Uganda. BMC Health Serv Res. 2015;15:347. doi: 10.1186/s12913-015-1019-5.

13. Warsame M, Gyapong M, Mpeka B, Rodrigues A, Singlovic J, Babiker A, et al. Pre-referral Rectal Artesunate Treatment by Community-Based Treatment Providers in Ghana, Guinea-Bissau, Tanzania, and Uganda (Study 18): A Cluster-Randomized Trial. Clin Infect Dis. 2016;63(Suppl 5):S312–S21. doi: 10.1093/cid/ciw631.

14. de Zoysa I, Bhandari N, Akhtari N, Bhan MK. Careseeking for illness in young infants in an urban slum in India. Soc Sci Med. 1998;47(12):2101–11. doi: 10.1016/s0277-9536(98)00275-5.

15. Kalter HD, Salgado R, Moulton LH, Nieto P, Contreras A, Egas ML, et al. Factors constraining adherence to referral advice for severely ill children managed by the Integrated Management of Childhood Illness approach in Imbabura Province, Ecuador. Acta Paediatr. 2003;92(1):103–10. doi: 10.1111/j.1651-2227.2003.tb00478.x.

16. Thomson A, Khogali M, de Smet M, Reid T, Mukhtar A, Peterson S, et al. Low referral completion of rapid diagnostic test-negative patients in community-based treatment of malaria in Sierra Leone. Malar J. 2011;10:94. doi: 10.1186/1475-2875-10-94.

17. Lengeler C, Burri C, Awor P, Athieno P, Kimera J, Tumukunde G, et al. Community access to rectal artesunate for malaria (CARAMAL): a large-scale observational implementation study in the Democratic Republic of the Congo, Nigeria and Uganda. medRxiv. 2021:2021.12.10.21266567. doi: 10.1101/2021.12.10.21266567.

18. Hetzel MW, Okitawutshu J, Tshefu A, Omoluabi E, Awor P, Signorell A, et al. Effectiveness of rectal artesunate as pre-referral treatment for severe malaria in children <5 years of age. medRxiv. 2021:2021.09.24.21263966. doi: 10.1101/2021.09.24.21263966.

19. Signorell A, Awor P, Okitawutshu J, Tshefu A, Omoluabi E, Hetzel MW, et al. Health worker compliance with severe malaria treatment guidelines in the context of implementing pre-referral rectal artesunate: an operational study in three high burden countries. medRxiv. 2021:2021.11.26.21266917. doi: 10.1101/2021.11.26.21266917.

20. Weiss DJ, Nelson A, Gibson HS, Temperley W, Peedell S, Lieber A, et al. A global map of travel time to cities to assess inequalities in accessibility in 2015. Nature. 2018;553(7688):333–6. doi: 10.1038/nature25181.

21. RStudio Team. RStudio: Integrated Development for R. Boston, MA: RStudio, PBC; 2020.

22. Bertozzi-Villa A. Mapping Travel Times with malariaAtlas and Friction Surfaces 2018 [19.05.2021]. Available from: https://medium.com/@abertozz/mapping-travel-times-with-malariaatlas-and-friction-surfaces-f4960f584f08.

23. Maina J, Ouma PO, Macharia PM, Alegana VA, Mitto B, Fall IS, et al. A spatial database of health facilities managed by the public health sector in sub Saharan Africa. Sci Data. 2019;6(1):134. doi: 10.1038/s41597-019-0142-2.

24. Google. [Google Maps referral health facility locations in the Democratic Republic of the Congo, Nigeria, and Uganda] n.d. Available from: https://www.google.com/maps.

25. Federal Ministry of Health Nigeria. NIGERIA Health Facility Registry n.d. Available from: https://hfr.health.gov.ng/facilities/hospitals-list.

26. Ministry of Health Uganda. National Health Facility Master List 2018 Kampala: Ministry of Health Uganda, Division of Health Information; 2018. Available from: http://library.health.go.ug/publications/health-facility-inventory/national-health-facility-master-facility-list-2018.

27. StataCorp. Stata Statistical Software: Release 16. College Station, TX: StataCorp LLC; 2019.

28. Awad MI, Alkadru AM, Behrens RH, Baraka OZ, Eltayeb IB. Descriptive study on the efficacy and safety of artesunate suppository in combination with other antimalarials in the treatment of severe malaria in Sudan. Am J Trop Med Hyg. 2003;68(2):153–8.

29. Barnes KI, Mwenechanya J, Tembo M, McIlleron H, Folb PI, Ribeiro I, et al. Efficacy of rectal artesunate compared with parenteral quinine in initial treatment of moderately severe malaria in African children and adults: a randomised study. Lancet. 2004;363(9421):1598–605. doi: 10.1016/S0140-6736(04)16203-X.

30. Krishna S, Planche T, Agbenyega T, Woodrow C, Agranoff D, Bedu-Addo G, et al. Bioavailability and preliminary clinical efficacy of intrarectal artesunate in Ghanaian children with moderate malaria. Antimicrob Agents Chemother. 2001;45(2):509–16. doi: 10.1128/AAC.45.2.509-516.2001.

31. Simpson JA, Agbenyega T, Barnes KI, Di Perri G, Folb P, Gomes M, et al. Population pharmacokinetics of artesunate and dihydroartemisinin following intra-rectal dosing of artesunate in malaria patients. PLoS Med. 2006;3(11):e444. doi: 10.1371/journal.pmed.0030444.

32. Dillip A, Alba S, Mshana C, Hetzel MW, Lengeler C, Mayumana I, et al. Acceptability--a neglected dimension of access to health care: findings from a study on childhood convulsions in rural Tanzania. BMC Health Serv Res. 2012;12:113. doi: 10.1186/1472-6963-12-113.

33. Hildenwall H, Tomson G, Kaija J, Pariyo G, Peterson S. “I never had the money for blood testing” - caretakers’ experiences of care-seeking for fatal childhood fevers in rural Uganda - a mixed methods study. BMC Int Health Hum Rights. 2008;8:12. doi: 10.1186/1472-698X-8-12.

34. Desmond NA, Nyirenda D, Dube Q, Mallewa M, Molyneux E, Lalloo DG, et al. Recognising and treatment seeking for acute bacterial meningitis in adults and children in resource-poor settings: a qualitative study. PLoS One. 2013;8(7):e68163. doi: 10.1371/journal.pone.0068163.

35. Brunner NC, Karim A, Athieno P, Kimera J, Tumukunde G, Angiro I, et al. Starting at the community: Treatment seeking pathways of children with suspected severe malaria in Uganda. medRxiv. 2021:2021.12.09.21267055. doi: 10.1101/2021.12.09.21267055.

36. Green C, Quigley P, Kureya T, Barber C, Chizema E, Moonga H, et al. Use of rectal artesunate for severe malaria at the community level, Zambia. Bull World Health Organ. 2019;97(12):810–7. doi: 10.2471/BLT.19.231506.

37. World Health Organization. Revised WHO classification and treatment of pneumonia in children at health facilities: evidence summary. Geneva: World Health Organization, 2014.

38. Lee TT, Omoluabi E, Ayodeji K, Yusuf O, Okon C, Brunner NC, et al. Treatment-seeking for children with suspected severe malaria attending community health workers and primary health centres in Adamawa State, Nigeria. medRxiv. 2021:2021.12.01.21267130. doi: 10.1101/2021.12.01.21267130.

39. Brunner N, Awor P, Hetzel M. Definitions of Severity in Treatment Seeking Studies of Febrile Illness in Children in Low and Middle Income Countries: A Scoping Review. Int J Public Health. 2021;66(74). doi: 10.3389/ijph.2021.634000.

