## Supplemental files for "Pre-referral rectal artesunate and referral completion among children with suspected severe malaria in the Democratic Republic of the Congo, Nigeria and Uganda"

### Supplementary material

### Supplement 1

**Inclusion flow-charts**

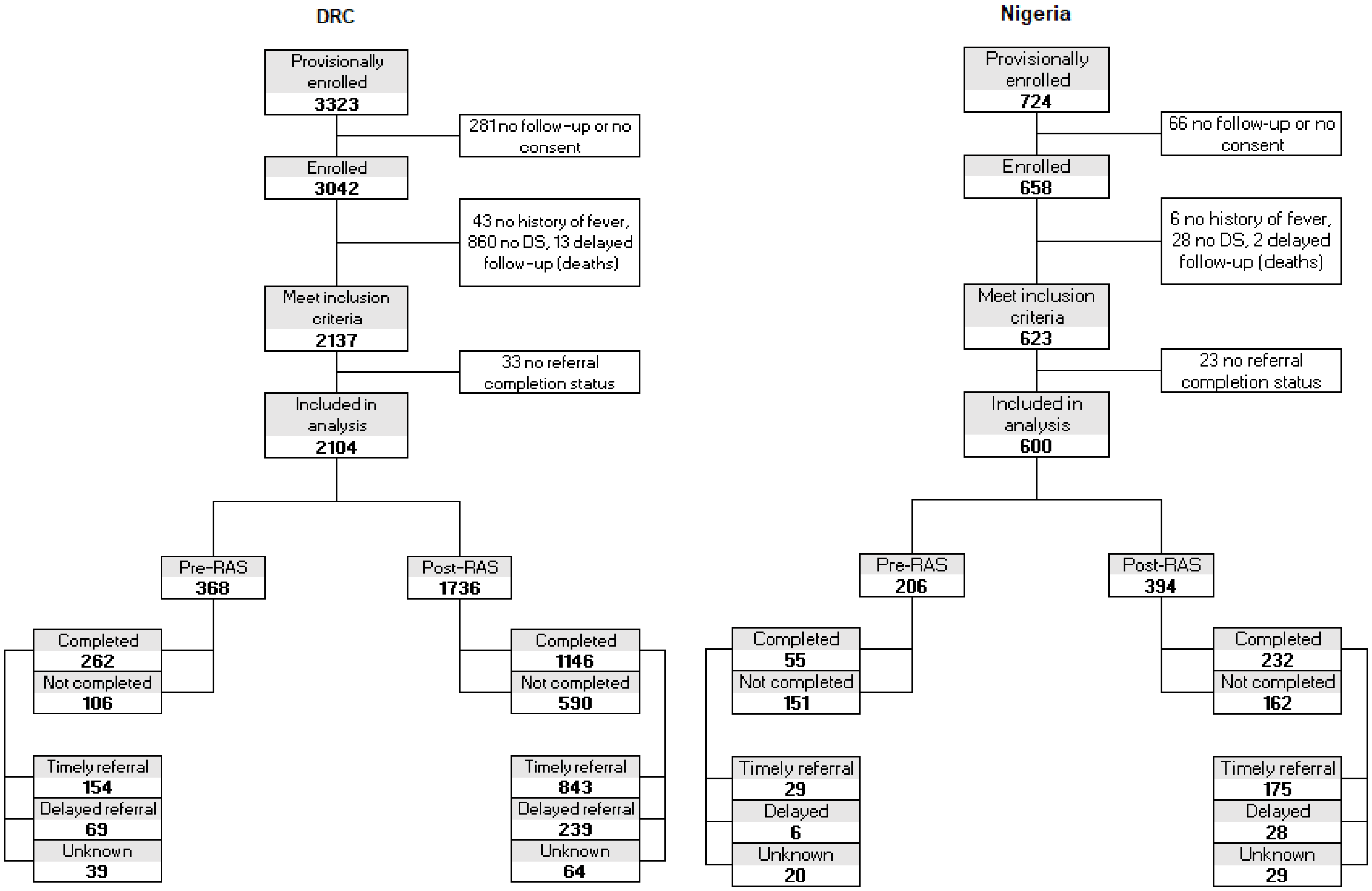

**Uganda**

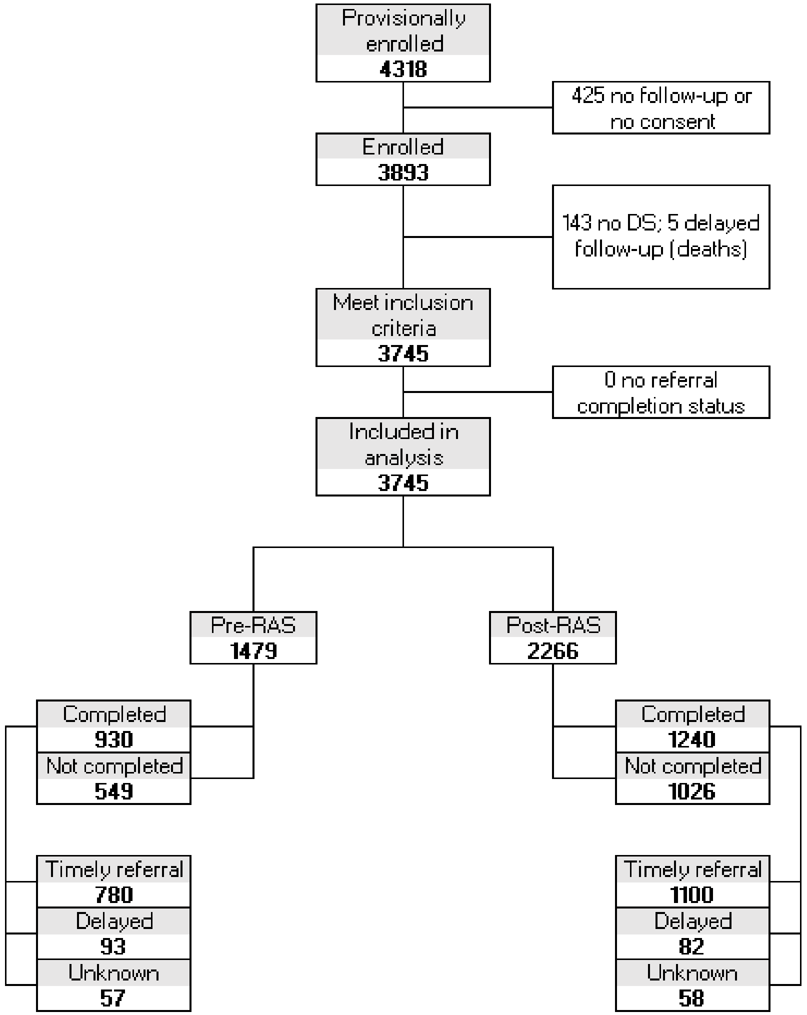
Provisionally enrolled
4318
425 no follow-up or no consent
Enrolled
3893
143 no DS; 5 delayed follow-up (deaths)
Meet inclusion criteria
3745
0 no referral completion status
Included in analysis
3745
Pre-RAS
1479
Post-RAS
2266
Completed
930
Not completed
549
Completed
1240
Not completed
1026
Timely referral
780
Delayed
93
Unknown
57
Timely referral
1100
Delayed
82
Unknown
58

### Supplement 2

**Table S2 1. Estimated associations between child characteristics and referral completion, DRC.**

| | DRC | | | | | | | | |
|---|---|---|---|---|---|---|---|---|---|
| | n | N | % | Unadjusted OR | 95% CI | p-value | Adjusted OR | 95% CI | p-value |
| **All** | **1,408** | **2,104** | **66.9** | | | | | | |
| **Study group** | | | | | | | | | |
| Pre-RAS | 262 | 368 | 71.2 | Ref. | | | Ref. | | |
| Post-RAS | | | | | | | | | |
| No RAS use | 142 | 188 | 75.5 | 0.42 | 0.23–0.77 | 0.005 | 0.34 | 0.18–0.66 | 0.001 |
| RAS use | 1,004 | 1,548 | 64.9 | 0.54 | 0.36–0.82 | 0.004 | 0.48 | 0.30–0.77 | 0.002 |
| **Child sex** | | | | | | | | | |
| Male | 735 | 1,121 | 65.6 | Ref. | | | Ref. | | |
| Female | 673 | 983 | 68.5 | 0.97 | 0.74–1.28 | 0.85 | 1.01 | 0.75–1.34 | 0.97 |
| **Age** | | | | | | | | | |
| 0 | 312 | 416 | 75.0 | Ref. | | | Ref. | | |
| 1 | 441 | 624 | 70.7 | 0.72 | 0.48–1.08 | 0.11 | 0.78 | 0.51–1.19 | 0.25 |
| 2 | 303 | 467 | 64.9 | 0.74 | 0.49–1.14 | 0.18 | 0.77 | 0.49–1.20 | 0.24 |
| 3 | 187 | 308 | 60.7 | 0.62 | 0.39–1.01 | 0.05 | 0.69 | 0.42–1.14 | 0.15 |
| 4 | 165 | 289 | 57.1 | 0.56 | 0.34–0.91 | 0.02 | 0.53 | 0.31–0.88 | 0.02 |
| **Caregiver age** | | | | | | | | | |
| <25 | 176 | 243 | 72.4 | Ref. | | | Ref. | | |
| 25–34 | 530 | 765 | 69.3 | 1.19 | 0.74–1.93 | 0.47 | 1.32 | 0.79–2.19 | 0.29 |
| 35–44 | 495 | 791 | 62.6 | 1.39 | 0.84–2.28 | 0.20 | 1.62 | 0.94–2.78 | 0.08 |
| >=45 | 207 | 305 | 67.9 | 1.28 | 0.73–2.27 | 0.39 | 1.38 | 0.74–2.57 | 0.31 |
| Missing | 0 | 0 | NA | NA | | | NA | | |
| **Caregiver sex** | | | | | | | | | |
| Male | 789 | 1,265 | 62.4 | Ref. | | | Ref. | | |
| Female | 619 | 839 | 73.8 | 1.10 | 0.81–1.49 | 0.54 | 1.19 | 0.85–1.67 | 0.31 |
| **Health zone** | | | | | | | | | |
| Ipamu | 562 | 692 | 81.2 | Ref. | | | Ref. | | |
| Kenge | 488 | 766 | 63.7 | 0.10 | 0.03–0.30 | <0.001 | 0.16 | 0.05–0.49 | 0.001 |
| Kingandu | 358 | 646 | 55.4 | 0.51 | 0.17–1.54 | 0.23 | 0.64 | 0.21–1.98 | 0.44 |
| **CNS involvement*** | | | | | | | | | |
| No | 532 | 678 | 78.5 | Ref. | | | Ref. | | |
| Yes | 876 | 1,426 | 61.4 | 0.60 | 0.43–0.83 | 0.002 | 0.58 | 0.41–0.82 | 0.002 |
| **Malaria test** | | | | | | | | | |
| Negative / Not done | 64 | 100 | 64.0 | Ref. | | | Ref. | | |
| Positive | 1,344 | 2,004 | 67.1 | 1.46 | 0.77–2.74 | 0.24 | 4.02 | 1.87–8.60 | <0.001 |
| **Enrolment location** | | | | | | | | | |
| CHW | 42 | 96 | 43.8 | Ref. | | | Ref. | | |
| PHC | 1,366 | 2,008 | 68.0 | 6.09 | 1.51–24.57 | 0.01 | 4.85 | 1.22–19.25 | 0.02 |

| | | | | | | | | | |
|---|---|---|---|---|---|---|---|---|---|
| **Enrolled during rainy season**** | | | | | | | | | |
| No | 665 | 959 | 69.3 | Ref. | | | Ref. | | |
| Yes | 743 | 1,145 | 64.9 | 0.97 | 0.73–1.28 | 0.82 | 0.77 | 0.56–1.07 | 0.12 |
| **Enrolled on a workday** | | | | | | | | | |
| No | 365 | 527 | 69.3 | Ref. | | | Ref. | | |
| Yes | 1,043 | 1,577 | 66.1 | 0.93 | 0.67–1.28 | 0.65 | 0.91 | 0.65–1.29 | 0.60 |
| **Enrolled during Covid-19 pandemic** | | | | | | | | | |
| No | 1,092 | 1,638 | 66.7 | Ref. | | | Ref. | | |
| Yes | 316 | 466 | 67.8 | 1.12 | 0.80–1.58 | 0.50 | 1.15 | 0.78–1.70 | 0.48 |
| **Delay to enrolling provider** | | | | | | | | | |
| 0–1 days | 402 | 667 | 60.3 | Ref. | | | Ref. | | |
| > 1 day | 915 | 1,336 | 68.5 | 1.33 | 0.98–1.79 | 0.06 | 1.07 | 0.77–1.49 | 0.69 |
| Missing | 91 | 101 | 90.1 | 7.87 | 3.08–20.10 | <0.001 | 3.33 | 0.86–12.83 | 0.08 |
| **Transport to enrolling provider** | | | | | | | | | |
| By foot / at home | 1,065 | 1,691 | 63.0 | Ref. | | | Ref. | | |
| Vehicle | 251 | 307 | 81.8 | 1.05 | 0.65–1.70 | 0.85 | 1.08 | 0.66–1.79 | 0.75 |
| Missing | 92 | 106 | 86.8 | 6.90 | 2.99–15.89 | <0.001 | 4.80 | 1.37–16.84 | 0.01 |
| **Time to RHF (min)** | | | | | | | | | |
| 0–<15 | 668 | 806 | 82.9 | Ref. | | | Ref. | | |
| 15–<30 | 232 | 322 | 72.0 | 1.01 | 0.60–1.71 | 0.98 | 1.12 | 0.65–1.95 | 0.68 |
| 30–<60 | 228 | 336 | 67.9 | 0.85 | 0.50–1.43 | 0.54 | 0.80 | 0.46–1.40 | 0.44 |
| ≥60 | 80 | 228 | 35.1 | 0.55 | 0.29–1.03 | 0.06 | 0.46 | 0.24–0.89 | 0.02 |
| Missing | 200 | 412 | 48.5 | 0.75 | 0.46–1.23 | 0.26 | 0.87 | 0.52–1.46 | 0.60 |
| **Perceived severity** | | | | | | | | | |
| Not fatal | 979 | 1,542 | 63.5 | Ref. | | | Ref. | | |
| Fatal | 413 | 545 | 75.8 | 1.57 | 1.10–2.25 | 0.01 | 1.86 | 1.28–2.71 | 0.001 |
| Missing | 16 | 17 | 94.1 | 33.29 | 1.39–795.57 | 0.03 | 16.14 | 0.61–429.60 | 0.10 |
| **Home treatment** | | | | | | | | | |
| No | 445 | 793 | 56.1 | Ref. | | | Ref. | | |
| Yes | 963 | 1,311 | 73.5 | 1.51 | 1.12–2.03 | 0.01 | 1.43 | 1.03–1.99 | 0.03 |
| Missing | 0 | 0 | NA | NA | | | NA | | |

* Danger signs involving the central nervous system (CNS): convulsions, unusually sleepy or unconscious

** October-April

**Table S2 2. Estimated associations between child characteristics and referral completion, Nigeria.**

| | Nigeria | | | | | | | | |
|---|---|---|---|---|---|---|---|---|---|
| | n | N | % | Unadjusted OR | 95% CI | p-value | Adjusted OR | 95% CI | p-value |
| **All** | **287** | **600** | **47.8** | | | | | | |
| **Study group** | | | | | | | | | |
| Pre-RAS | 55 | 206 | 26.7 | Ref. | | | NA | | |
| Post-RAS | | | | | | | | | |
| No RAS use | 118 | 183 | 64.5 | 5.95 | 2.54–13.93 | <0.001 | NA | | |
| RAS use | 114 | 211 | 54.0 | 2.17 | 0.94–5.01 | 0.07 | NA | | |
| **Child sex** | | | | | | | | | |
| Male | 177 | 364 | 48.6 | Ref. | | | Ref. | | |
| Female | 110 | 236 | 46.6 | 0.67 | 0.37–1.23 | 0.20 | 0.58 | 0.29–1.17 | 0.13 |
| **Age** | | | | | | | | | |
| 0 | 28 | 71 | 39.4 | Ref. | | | Ref. | | |
| 1 | 85 | 163 | 52.1 | 1.51 | 0.51–4.47 | 0.46 | 1.88 | 0.57–6.29 | 0.30 |
| 2 | 86 | 170 | 50.6 | 1.07 | 0.37–3.10 | 0.90 | 1.23 | 0.38–3.92 | 0.73 |
| 3 | 50 | 119 | 42.0 | 0.63 | 0.21–1.92 | 0.42 | 0.74 | 0.21–2.59 | 0.64 |
| 4 | 38 | 77 | 49.4 | 1.57 | 0.45–5.45 | 0.47 | 1.96 | 0.50–7.64 | 0.33 |
| **Caregiver age** | | | | | | | | | |
| <25 | 52 | 93 | 55.9 | Ref. | | | Ref. | | |
| 25–34 | 140 | 300 | 46.7 | 0.53 | 0.21–1.34 | 0.18 | 0.55 | 0.19–1.59 | 0.27 |
| 35–44 | 68 | 149 | 45.6 | 0.43 | 0.16–1.18 | 0.10 | 0.53 | 0.16–1.75 | 0.30 |
| >=45 | 26 | 57 | 45.6 | 0.39 | 0.11–1.37 | 0.14 | 0.58 | 0.14–2.50 | 0.47 |
| Missing | 1 | 1 | 100.0 | NA | | | NA | | |
| **Caregiver sex** | | | | | | | | | |
| Male | 96 | 201 | 47.8 | Ref. | | | Ref. | | |
| Female | 191 | 399 | 47.9 | 1.88 | 1.00–3.54 | 0.05 | 1.35 | 0.61–3.00 | 0.46 |
| **LGA** | | | | | | | | | |
| Mayo-Belwa | 160 | 251 | 63.7 | Ref. | | | | | |
| Fufore | 84 | 245 | 34.3 | 0.33 | 0.07–1.49 | 0.15 | NA | | |
| Song | 43 | 104 | 41.3 | 1.00 | 0.19–5.28 | 1.00 | NA | | |
| **CNS involvement** | | | | | | | | | |
| No | 33 | 99 | 33.3 | Ref. | | | Ref. | | |
| Yes | 254 | 501 | 50.7 | 0.77 | 0.33–1.83 | 0.56 | 0.48 | 0.17–1.38 | 0.17 |
| **Malaria test** | | | | | | | | | |
| Negative / Not done | 15 | 32 | 46.9 | Ref. | | | Ref. | | |
| Positive | 272 | 568 | 47.9 | 1.45 | 0.35–5.94 | 0.61 | 4.31 | 0.69–26.75 | 0.12 |
| **Enrolment location** | | | | | | | | | |
| CHW | 49 | 315 | 15.6 | Ref. | | | NA | | |
| PHC | 238 | 285 | 83.5 | 85.47 | 23.16–315.38 | <0.001 | NA | | |
| **Enrolled during rainy season**** | | | | | | | | | |
| No | 78 | 154 | 50.6 | Ref. | | | Ref. | | |
| Yes | 209 | 446 | 46.9 | 0.64 | 0.32–1.29 | 0.21 | 0.70 | 0.29–1.67 | 0.42 |

| | | | | | | | | | |
|---|---|---|---|---|---|---|---|---|---|
| **Enrolled on a workday** | | | | | | | | | |
| No | 33 | 107 | 30.8 | Ref. | | | Ref. | | |
| Yes | 254 | 493 | 51.5 | 1.48 | 0.67–3.27 | 0.33 | 0.74 | 0.31–1.76 | 0.50 |
| **Enrolled during Covid-19 pandemic** | | | | | | | | | |
| No | 239 | 483 | 49.5 | Ref. | | | Ref. | | |
| Yes | 48 | 117 | 41.0 | 0.19 | 0.08–0.42 | <0.001 | 0.09 | 0.03–0.26 | <0.001 |
| **Delay to enrolling provider** | | | | | | | | | |
| 0–1 days | 90 | 197 | 45.7 | Ref. | | | Ref. | | |
| > 1 day | 154 | 327 | 47.1 | 1.35 | 0.71–2.58 | 0.36 | 1.64 | 0.77–3.52 | 0.20 |
| Missing | 43 | 76 | 56.6 | 2.27 | 0.80–6.44 | 0.12 | 3.53 | 1.03–12.09 | 0.04 |
| **Transport to enrolling provider** | | | | | | | | | |
| No vehicle / missing | 125 | 371 | 33.7 | Ref. | | | Ref. | | |
| Vehicle | 162 | 229 | 70.7 | 1.28 | 0.60–2.69 | 0.52 | 0.82 | 0.32–2.10 | 0.68 |
| **Time to RHF (min)** | | | | | | | | | |
| 0–<15 | 85 | 104 | 81.7 | Ref. | | | Ref. | | |
| 15–<30 | 62 | 87 | 71.3 | 0.60 | 0.17–2.14 | 0.43 | 0.48 | 0.13–1.73 | 0.26 |
| 30–<60 | 83 | 149 | 55.7 | 0.39 | 0.13–1.21 | 0.10 | 0.23 | 0.07–0.77 | 0.02 |
| > 60 | 48 | 247 | 19.4 | 0.13 | 0.04–0.43 | <0.001 | 0.06 | 0.02–0.22 | <0.001 |
| Missing | 9 | 13 | 69.2 | 0.85 | 0.10–7.15 | 0.88 | 0.13 | 0.01–1.84 | 0.13 |
| **Perceived severity** | | | | | | | | | |
| Not fatal | 212 | 434 | 48.8 | Ref. | | | Ref. | | |
| Fatal | 72 | 161 | 44.7 | 0.61 | 0.32–1.17 | 0.14 | 0.63 | 0.30–1.32 | 0.23 |
| Missing | 3 | 5 | 60.0 | 1.25 | 0.05–32.21 | 0.89 | 1.23 | 0.01–153.00 | 0.93 |
| **Home treatment** | | | | | | | | | |
| No / missing | 166 | 379 | 43.8 | Ref. | | | Ref. | | |
| Yes | 121 | 221 | 54.8 | 1.14 | 0.63–2.06 | 0.68 | 1.08 | 0.54–2.16 | 0.82 |
| **Interactions** | | | | | | | | | |
| **Enrolment location (adjusted for study group)** | | | | | | | | | |
| CHW | 49 | 315 | 15.6 | Ref. | | | Ref. | | |
| PHC | 238 | 285 | 83.5 | 92.82 | 18.92–455.42 | <0.001 | 19.79 | 2.97–131.71 | 0.002 |
| **Study group by enrolment location** | | | | | | | | | |
| **CHW** | | | | | | | | | |
| Pre-RAS | 9 | 148 | 6 | Ref. | | | Ref. | | |
| Post-RAS | | | | | | | | | |
| No RAS use | 16 | 76 | 21 | 3.14 | 1.03–9.60 | 0.04 | 3.97 | 1.07–14.75 | 0.04 |
| RAS use | 24 | 91 | 26 | 5.33 | 1.75–16.20 | 0.003 | 9.95 | 2.71–36.58 | <0.001 |
| **PHC** | | | | | | | | | |
| Pre-RAS | 46 | 58 | 79 | Ref. | | | Ref. | | |
| Post-RAS | | | | | | | | | |
| No RAS use | 102 | 107 | 95 | 13.00 | 3.11–54.42 | <0.001 | 35.09 | 6.52–188.75 | <0.001 |
| RAS use | 90 | 120 | 75 | 1.23 | 0.43–3.50 | 0.70 | 6.45 | 1.62–25.67 | 0.01 |

| | | | | | | | | | |
|---|---|---|---|---|---|---|---|---|---|
| **Enrolment location by LGA** | | | | | | | | | |
| Mayo-Belwa - CHW | 26 | 95 | 27.4 | Ref. | | | Ref. | | |
| Mayo-Belwa - PHC | 134 | 156 | 85.9 | 16.62 | 3.50–78.93 | <0.001 | NA | | |
| Fufore - CHW | 5 | 145 | 3.4 | 0.04 | 0.01–0.25 | <0.001 | 0.07 | 0.01–0.43 | 0.004 |
| Fufore - PHC | 79 | 100 | 79.0 | 32.81 | 5.27–204.46 | <0.001 | 1.98 | 0.33–11.99 | 0.46 |
| Song - CHW | 18 | 75 | 24.0 | 0.62 | 0.15–2.58 | 0.51 | 0.50 | 0.12–2.07 | 0.34 |
| Song - PHC | 25 | 29 | 86.2 | 39.60 | 4.61–340.41 | <0.001 | 3.20 | 0.33–30.93 | 0.32 |

* Danger signs involving the central nervous system (CNS): convulsions, unusually sleepy or unconscious

** Mai-October

**Table S2 3. Estimated associations between child characteristics and referral completion, Uganda.**

| | Uganda | | | | | | | | |
|---|---|---|---|---|---|---|---|---|---|
| | n | N | % | Unadjusted OR | 95% CI | p-value | Adjusted OR | 95% CI | p-value |
| **All** | **2,170** | **3,745** | **57.9** | | | | | | |
| **Study group** | | | | | | | | | |
| Pre-RAS | 930 | 1,479 | 62.9 | Ref. | | | Ref. | | |
| Post-RAS | | | | | | | | | |
| No RAS use | 365 | 635 | 57.5 | 0.79 | 0.63–1.00 | 0.05 | 0.80 | 0.63–1.01 | 0.06 |
| RAS use | 875 | 1,631 | 53.6 | 0.64 | 0.53–0.77 | <0.001 | 0.72 | 0.58–0.88 | 0.002 |
| **Child sex** | | | | | | | | | |
| Male | 1,162 | 1,993 | 58.3 | Ref. | | | Ref. | | |
| Female | 1,008 | 1,752 | 57.5 | 0.97 | 0.83–1.12 | 0.66 | 0.98 | 0.84–1.14 | 0.81 |
| **Age** | | | | | | | | | |
| 0 | 417 | 675 | 61.8 | Ref. | | | Ref. | | |
| 1 | 637 | 1,078 | 59.1 | 0.92 | 0.74–1.15 | 0.48 | 0.93 | 0.74–1.17 | 0.54 |
| 2 | 500 | 890 | 56.2 | 0.74 | 0.59–0.93 | 0.01 | 0.74 | 0.58–0.94 | 0.01 |
| 3 | 401 | 694 | 57.8 | 0.84 | 0.65–1.07 | 0.15 | 0.83 | 0.65–1.07 | 0.16 |
| 4 | 215 | 408 | 52.7 | 0.69 | 0.52–0.92 | 0.01 | 0.69 | 0.51–0.92 | 0.01 |
| **Caregiver age** | | | | | | | | | |
| <25 | 831 | 1,394 | 59.6 | Ref. | | | Ref. | | |
| 25–34 | 868 | 1,484 | 58.5 | 0.92 | 0.78–1.09 | 0.36 | 0.93 | 0.78–1.11 | 0.44 |
| 35–44 | 327 | 608 | 53.8 | 0.82 | 0.66–1.03 | 0.08 | 0.84 | 0.66–1.06 | 0.13 |
| >=45 | 143 | 258 | 55.4 | 0.87 | 0.64–1.18 | 0.37 | 0.91 | 0.66–1.25 | 0.55 |
| Missing | 1 | 1 | 100.0 | NA | | | NA | | |
| **Caregiver sex** | | | | | | | | | |
| Male | 359 | 610 | 58.9 | Ref. | | | Ref. | | |
| Female | 1,811 | 3,135 | 57.8 | 0.96 | 0.78–1.18 | 0.70 | 0.92 | 0.74–1.14 | 0.46 |
| **Health zone** | | | | | | | | | |
| Ipamu | 1,071 | 1,749 | 61.2 | Ref. | | | Ref. | | |
| Kenge | 525 | 991 | 53.0 | 0.74 | 0.44–1.26 | 0.27 | 0.88 | 0.52–1.49 | 0.64 |
| Kingandu | 574 | 1,005 | 57.1 | 1.25 | 0.70–2.25 | 0.46 | 1.46 | 0.80–2.63 | 0.21 |
| **CNS involvement** | | | | | | | | | |
| No | 267 | 425 | 62.8 | Ref. | | | Ref. | | |
| Yes | 1,903 | 3,320 | 57.3 | 0.67 | 0.53–0.86 | 0.002 | 0.80 | 0.61–1.04 | 0.09 |
| **Malaria test** | | | | | | | | | |
| Negative / Not done | 40 | 59 | 67.8 | Ref. | | | Ref. | | |
| Positive | 2,130 | 3,686 | 57.8 | 0.81 | 0.43–1.49 | 0.49 | 0.90 | 0.48–1.67 | 0.74 |
| **Enrolment location** | | | | | | | | | |
| CHW | 2,170 | 3,745 | 57.9 | NA | | | NA | | |
| PHC | 0 | 0 | NA | NA | | | NA | | |
| **Enrolled during rainy season**** | | | | | | | | | |
| No | 772 | 1,392 | 55.5 | Ref. | | | Ref. | | |
| Yes | 1,398 | 2,353 | 59.4 | 1.15 | 0.99–1.35 | 0.07 | 1.15 | 0.97–1.38 | 0.11 |

| | | | | | | | | | |
|---|---|---|---|---|---|---|---|---|---|
| **Enrolled on a workday** | | | | | | | | | |
| No | 516 | 959 | 53.8 | Ref. | | | Ref. | | |
| Yes | 1,654 | 2,786 | 59.4 | 1.20 | 1.01–1.42 | 0.04 | 1.19 | 1.00–1.41 | 0.05 |
| **Enrolled during Covid-19 pandemic** | | | | | | | | | |
| No | 1,922 | 3,323 | 57.8 | Ref. | | | Ref. | | |
| Yes | 248 | 422 | 58.8 | 0.87 | 0.68–1.11 | 0.25 | 0.90 | 0.69–1.19 | 0.48 |
| **Delay to enrolling provider** | | | | | | | | | |
| 0–1 days | 1,212 | 2,134 | 56.8 | Ref. | | | Ref. | | |
| > 1 day | 925 | 1,565 | 59.1 | 1.15 | 0.99–1.35 | 0.08 | 1.14 | 0.97–1.34 | 0.12 |
| Missing | 33 | 46 | 71.7 | 1.87 | 0.89–3.94 | 0.10 | 1.11 | 0.43–2.86 | 0.83 |
| **Transport to enrolling provider** | | | | | | | | | |
| By foot / at home | 2,002 | 3,462 | 57.8 | Ref. | | | Ref. | | |
| Vehicle | 150 | 260 | 57.7 | 1.09 | 0.81–1.45 | 0.57 | 1.06 | 0.78–1.42 | 0.72 |
| Missing | 18 | 23 | 78.3 | 3.39 | 1.08–10.66 | 0.04 | 3.31 | 0.78–14.06 | 0.11 |
| **Time to RHF (min)** | | | | | | | | | |
| 0–<15 | 1,349 | 1,998 | 67.5 | Ref. | | | Ref. | | |
| 15–<30 | 692 | 1,457 | 47.5 | 0.69 | 0.56–0.85 | <0.001 | 0.72 | 0.58–0.89 | 0.003 |
| 30–<60 | 115 | 270 | 42.6 | 0.54 | 0.38–0.77 | <0.001 | 0.55 | 0.39–0.79 | 0.001 |
| > 60 | 0 | 2 | 0.0 | NA | | | NA | | |
| Missing | 14 | 18 | 77.8 | 2.35 | 0.68–8.14 | 0.18 | 1.88 | 0.53–6.67 | 0.33 |
| **Perceived severity** | | | | | | | | | |
| Not fatal | 1,180 | 2,078 | 56.8 | Ref. | | | Ref. | | |
| Fatal | 985 | 1,657 | 59.4 | 1.07 | 0.91–1.24 | 0.41 | 1.11 | 0.94–1.30 | 0.21 |
| Missing | 5 | 10 | 50.0 | 0.78 | 0.20–3.15 | 0.73 | 0.80 | 0.20–3.19 | 0.75 |
| **Home treatment** | | | | | | | | | |
| No | 1,876 | 3,271 | 57.4 | Ref. | | | Ref. | | |
| Yes | 294 | 474 | 62.0 | 1.18 | 0.94–1.49 | 0.15 | 1.10 | 0.87–1.39 | 0.41 |
| Missing | 0 | 0 | NA | NA | | | NA | | |

* Danger signs involving the central nervous system (CNS): convulsions, unusually sleepy or unconscious

** April-October

### Supplement 3

**Table S3 1. Estimated associations between child characteristics and referral timeliness, DRC.**

| | DRC | | | | | | | | |
|---|---|---|---|---|---|---|---|---|---|
| | n | N | % | Undadjusted OR | 95% CI | p-value | Adjusted OR | 95% CI | p-value |
| **All** | **997** | **1,305** | **76.4** | | | | | | |
| **Study group** | | | | | | | | | |
| Pre-RAS | 154 | 223 | 69.1 | Ref. | | | Ref. | | |
| Post-RAS | | | | | | | | | |
| No RAS use | 89 | 128 | 69.5 | 1.01 | 0.61–1.69 | 0.96 | 0.92 | 0.52–1.62 | 0.77 |
| RAS use | 754 | 954 | 79.0 | 1.49 | 1.04–2.14 | 0.03 | 1.39 | 0.92–2.12 | 0.12 |
| **Child sex** | | | | | | | | | |
| Male | 513 | 680 | 75.4 | Ref. | | | Ref. | | |
| Female | 484 | 625 | 77.4 | 1.08 | 0.82–1.42 | 0.58 | 1.11 | 0.83–1.48 | 0.47 |
| **Age** | | | | | | | | | |
| 0 | 207 | 285 | 72.6 | Ref. | | | Ref. | | |
| 1 | 323 | 413 | 78.2 | 1.46 | 1.00–2.13 | 0.05 | 1.56 | 1.06–2.31 | 0.02 |
| 2 | 214 | 283 | 75.6 | 1.29 | 0.86–1.95 | 0.21 | 1.26 | 0.83–1.92 | 0.28 |
| 3 | 140 | 173 | 80.9 | 1.71 | 1.03–2.82 | 0.04 | 1.48 | 0.88–2.49 | 0.14 |
| 4 | 113 | 151 | 74.8 | 1.09 | 0.66–1.82 | 0.73 | 1.07 | 0.63–1.81 | 0.80 |
| **Caregiver age** | | | | | | | | | |
| <25 | 124 | 164 | 75.6 | Ref. | | | Ref. | | |
| 25–34 | 377 | 500 | 75.4 | 0.96 | 0.62–1.50 | 0.87 | 0.85 | 0.53–1.35 | 0.49 |
| 35–44 | 357 | 458 | 77.9 | 1.01 | 0.64–1.61 | 0.95 | 0.85 | 0.52–1.40 | 0.53 |
| >=45 | 139 | 183 | 76.0 | 0.83 | 0.48–1.41 | 0.48 | 0.76 | 0.42–1.34 | 0.34 |
| Not documented | 0 | 0 | NA | NA | | | NA | | |
| **Caregiver sex** | | | | | | | | | |
| Male | 557 | 716 | 77.8 | Ref. | | | Ref. | | |
| Female | 440 | 589 | 74.7 | 0.86 | 0.63–1.16 | 0.31 | 0.78 | 0.56–1.09 | 0.15 |
| **Health zone** | | | | | | | | | |
| Ipamu | 418 | 514 | 81.3 | Ref. | | | Ref. | | |
| Kenge | 345 | 485 | 71.1 | 0.53 | 0.29–0.97 | 0.04 | 0.54 | 0.29–1.02 | 0.06 |
| Kingandu | 234 | 306 | 76.5 | 0.72 | 0.38–1.34 | 0.29 | 0.75 | 0.38–1.49 | 0.41 |
| **CNS involvement*** | | | | | | | | | |
| No | 358 | 493 | 72.6 | Ref. | | | Ref. | | |
| Yes | 639 | 812 | 78.7 | 1.55 | 1.16–2.07 | 0.003 | 1.50 | 1.10–2.04 | 0.01 |
| **Malaria test** | | | | | | | | | |
| Negative / Not done | 38 | 47 | 80.9 | Ref. | | | Ref. | | |
| Positive | 959 | 1,258 | 76.2 | 0.71 | 0.31–1.63 | 0.41 | 0.39 | 0.13–1.21 | 0.10 |
| **Enrolment location** | | | | | | | | | |
| CHW | 31 | 38 | 81.6 | Ref. | | | Ref. | | |
| PHC | 966 | 1,267 | 76.2 | 0.65 | 0.22–1.92 | 0.43 | 0.63 | 0.21–1.86 | 0.40 |

| | | | | | | | | | |
|---|---|---|---|---|---|---|---|---|---|
| **Enrolled during rainy season**** | | | | | | | | | |
| No | 501 | 627 | 79.9 | Ref. | | | Ref. | | |
| Yes | 496 | 678 | 73.2 | 0.71 | 0.54–0.95 | 0.02 | 0.73 | 0.53–1.01 | 0.06 |
| **Enrolled on a workday** | | | | | | | | | |
| No | 267 | 341 | 78.3 | Ref. | | | Ref. | | |
| Yes | 730 | 964 | 75.7 | 0.83 | 0.61–1.15 | 0.26 | 0.83 | 0.59–1.15 | 0.25 |
| **Enrolled during Covid-19 pandemic** | | | | | | | | | |
| No | 764 | 1,005 | 76.0 | Ref. | | | Ref. | | |
| Yes | 233 | 300 | 77.7 | 1.09 | 0.77–1.54 | 0.62 | 0.83 | 0.57–1.22 | 0.35 |
| **Delay to enrolling provider** | | | | | | | | | |
| 0–1 days | 296 | 396 | 74.7 | Ref. | | | Ref. | | |
| > 1 day | 671 | 873 | 76.9 | 1.19 | 0.87–1.61 | 0.27 | 1.69 | 1.20–2.38 | 0.003 |
| Not documented | 30 | 36 | 83.3 | 1.43 | 0.51–4.01 | 0.50 | 2.29 | 0.35–14.86 | 0.38 |
| **Transport to enrolling provider** | | | | | | | | | |
| By foot / at home | 786 | 1,024 | 76.8 | Ref. | | | Ref. | | |
| Vehicle | 181 | 243 | 74.5 | 0.82 | 0.56–1.21 | 0.33 | 0.88 | 0.59–1.31 | 0.53 |
| Not documented | 30 | 38 | 78.9 | 0.93 | 0.37–2.34 | 0.88 | 0.37 | 0.07–1.97 | 0.25 |
| **Time to RHF (min)** | | | | | | | | | |
| 0–<15 | 498 | 636 | 78.3 | Ref. | | | Ref. | | |
| 15–<30 | 157 | 213 | 73.7 | 0.95 | 0.59–1.53 | 0.84 | 0.98 | 0.59–1.61 | 0.92 |
| 30–<60 | 147 | 202 | 72.8 | 0.83 | 0.51–1.35 | 0.46 | 0.82 | 0.49–1.38 | 0.46 |
| ≥60 | 44 | 69 | 63.8 | 0.60 | 0.31–1.16 | 0.13 | 0.58 | 0.28–1.18 | 0.13 |
| Missing | 151 | 185 | 81.6 | 1.32 | 0.80–2.17 | 0.28 | 1.25 | 0.72–2.15 | 0.43 |
| **Perceived severity** | | | | | | | | | |
| Not fatal | 690 | 907 | 76.1 | Ref. | | | Ref. | | |
| Fatal | 294 | 384 | 76.6 | 0.91 | 0.67–1.26 | 0.59 | 0.92 | 0.66–1.28 | 0.61 |
| Not documented | 13 | 14 | 92.9 | 1.63 | 0.18–14.28 | 0.66 | 1.73 | 0.19–15.97 | 0.63 |
| **Home treatment** | | | | | | | | | |
| No | 339 | 406 | 83.5 | Ref. | | | Ref. | | |
| Yes | 658 | 899 | 73.2 | 0.53 | 0.38–0.75 | <0.001 | 0.49 | 0.33–0.71 | <0.001 |
| Not documented | 0 | 0 | NA | NA | | | NA | | |

* Danger sign involving the central nervous system (CNS): convulsions, unusually sleepy or unconscious

** October-April

**Table S3 2. Estimated associations between child characteristics and referral timeliness, Nigeria.**

| | Nigeria | | | | | | | | |
|---|---|---|---|---|---|---|---|---|---|
| | n | N | % | Undadjusted OR | 95% CI | p-value | Adjusted OR | 95% CI | p-value |
| **All** | **204** | **238** | **85.7** | | | | | | |
| **Study group** | | | | | | | | | |
| Pre-RAS | 29 | 35 | 82.9 | Ref. | | | Ref. | | |
| Post-RAS | | | | | | | | | |
| No RAS use | 84 | 101 | 83.2 | 1.02 | 0.37–2.84 | 0.97 | 1.33 | 0.36–4.85 | 0.67 |
| RAS use | 91 | 102 | 89.2 | 1.71 | 0.58–5.04 | 0.33 | 1.53 | 0.36–6.40 | 0.56 |
| **Child sex** | | | | | | | | | |
| Male | 114 | 140 | 81.4 | Ref. | | | Ref. | | |
| Female | 90 | 98 | 91.8 | 2.57 | 1.11–5.94 | 0.03 | 2.73 | 1.03–7.28 | 0.04 |
| **Age** | | | | | | | | | |
| 0 | 21 | 24 | 87.5 | Ref. | | | Ref. | | |
| 1 | 58 | 70 | 82.9 | 0.69 | 0.18–2.69 | 0.59 | 0.48 | 0.09–2.67 | 0.40 |
| 2 | 64 | 73 | 87.7 | 1.02 | 0.25–4.11 | 0.98 | 0.68 | 0.13–3.65 | 0.65 |
| 3 | 35 | 39 | 89.7 | 1.25 | 0.25–6.14 | 0.78 | 0.83 | 0.12–5.87 | 0.86 |
| 4 | 26 | 32 | 81.3 | 0.62 | 0.14–2.78 | 0.53 | 0.54 | 0.08–3.54 | 0.52 |
| **Caregiver age** | | | | | | | | | |
| <25 | 42 | 48 | 87.5 | Ref. | | | Ref. | | |
| 25–34 | 98 | 117 | 83.8 | 0.74 | 0.27–1.98 | 0.54 | 0.40 | 0.12–1.35 | 0.14 |
| 35–44 | 48 | 54 | 88.9 | 1.14 | 0.34–3.81 | 0.83 | 0.49 | 0.11–2.12 | 0.34 |
| >=45 | 16 | 19 | 84.2 | 0.76 | 0.17–3.42 | 0.72 | 0.25 | 0.04–1.69 | 0.15 |
| Not documented | 0 | 0 | NA | NA | | | NA | | |
| **Caregiver sex** | | | | | | | | | |
| Male | 63 | 69 | 91.3 | Ref. | | | Ref. | | |
| Female | 141 | 169 | 83.4 | 0.48 | 0.19–1.22 | 0.12 | 0.42 | 0.12–1.41 | 0.16 |
| **LGA** | | | | | | | | | |
| Mayo-Belwa | 112 | 135 | 83.0 | Ref. | | | Ref. | | |
| Fufore | 56 | 65 | 86.2 | 1.31 | 0.48–3.56 | 0.60 | 0.84 | 0.30–2.40 | 0.75 |
| Song | 36 | 38 | 94.7 | 3.78 | 0.78–18.32 | 0.10 | 5.42 | 1.02–28.68 | 0.05 |
| **CNS involvement** | | | | | | | | | |
| No | 18 | 26 | 69.2 | Ref. | | | Ref. | | |
| Yes | 186 | 212 | 87.7 | 3.18 | 1.26–8.05 | 0.01 | 4.78 | 1.28–17.77 | 0.02 |
| **Malaria test** | | | | | | | | | |
| Negative / Not done | 9 | 10 | 90.0 | Ref. | | | Ref. | | |
| Positive | 195 | 228 | 85.5 | 0.66 | 0.08–5.35 | 0.69 | 1.58 | 0.15–16.73 | 0.71 |
| **Enrolment location** | | | | | | | | | |
| CHW | 30 | 39 | 76.9 | Ref. | | | Ref. | | |
| PHC | 174 | 199 | 87.4 | 2.09 | 0.89–4.91 | 0.09 | 1.73 | 0.45–6.60 | 0.42 |
| **Enrolled during rainy season**** | | | | | | | | | |
| No | 56 | 66 | 84.8 | Ref. | | | Ref. | | |
| Yes | 148 | 172 | 86.0 | 1.10 | 0.50–2.45 | 0.81 | 0.59 | 0.20–1.74 | 0.34 |

| | | | | | | | | | |
|---|---|---|---|---|---|---|---|---|---|
| **Enrolled on a workday** | | | | | | | | | |
| No | 24 | 27 | 88.9 | Ref. | | | Ref. | | |
| Yes | 180 | 211 | 85.3 | 0.73 | 0.21–2.56 | 0.62 | 0.78 | 0.18–3.41 | 0.74 |
| **Enrolled during Covid-19 pandemic** | | | | | | | | | |
| No | 165 | 195 | 84.6 | Ref. | | | Ref. | | |
| Yes | 39 | 43 | 90.7 | 1.77 | 0.59–5.33 | 0.31 | 1.90 | 0.49–7.44 | 0.35 |
| **Delay to enrolling provider** | | | | | | | | | |
| 0–1 days | 77 | 89 | 86.5 | Ref. | | | Ref. | | |
| > 1 day | 122 | 144 | 84.7 | 0.86 | 0.40–1.85 | 0.71 | 0.63 | 0.25–1.58 | 0.33 |
| Not documented | 5 | 5 | 100.0 | NA | | | NA | | |
| **Transport to enrolling provider** | | | | | | | | | |
| No vehicle | 70 | 84 | 83.3 | Ref. | | | Ref. | | |
| Vehicle | 134 | 154 | 87.0 | 1.44 | 0.69–3.04 | 0.33 | 1.14 | 0.40–3.24 | 0.81 |
| Not documented | 5 | 5 | 100.0 | NA | | | NA | | |
| **Time to RHF (min)** | | | | | | | | | |
| 0–<15 | 62 | 70 | 88.6 | Ref. | | | Ref. | | |
| 15–<30 | 46 | 48 | 95.8 | 2.97 | 0.60–14.64 | 0.18 | 1.72 | 0.31–9.69 | 0.54 |
| 30–<60 | 59 | 75 | 78.7 | 0.48 | 0.19–1.19 | 0.11 | 0.38 | 0.14–1.06 | 0.07 |
| > 60 | 34 | 41 | 82.9 | 0.63 | 0.21–1.88 | 0.40 | 0.75 | 0.20–2.83 | 0.67 |
| Missing | 3 | 4 | 75.0 | 0.39 | 0.04–4.18 | 0.43 | 0.16 | 0.01–2.76 | 0.21 |
| **Perceived severity** | | | | | | | | | |
| Not fatal | 155 | 180 | 86.1 | Ref. | | | Ref. | | |
| Fatal | 49 | 58 | 84.5 | 0.88 | 0.38–2.01 | 0.76 | 0.93 | 0.34–2.52 | 0.88 |
| Not documented | 0 | 0 | NA | NA | | | NA | | |
| **Home treatment** | | | | | | | | | |
| No | 117 | 138 | 84.8 | Ref. | | | Ref. | | |
| Yes | 87 | 100 | 87.0 | 1.20 | 0.57–2.53 | 0.63 | 1.02 | 0.42–2.49 | 0.96 |
| Not documented | 0 | 0 | NA | NA | | | NA | | |

* Danger sign involving the central nervous system (CNS): convulsions, unusually sleepy or unconscious
** Mai-October

**Table S3 3. Estimated associations between child characteristics and referral timeliness, Uganda.**

| | Uganda | | | | | | | | |
|---|---|---|---|---|---|---|---|---|---|
| | n | N | % | Unadjusted OR | 95% CI | p-value | Adjusted OR | 95% CI | p-value |
| **All** | **1,880** | **2,055** | **91.5** | | | | | | |
| **Study group** | | | | | | | | | |
| Pre-RAS | 780 | 873 | 89.3 | Ref. | | | Ref. | | |
| Post-RAS | | | | | | | | | |
| No RAS use | 321 | 350 | 91.7 | 1.40 | 0.88–2.20 | 0.15 | 1.43 | 0.89–2.30 | 0.14 |
| RAS use | 779 | 832 | 93.6 | 1.91 | 1.31–2.79 | <0.001 | 1.81 | 1.17–2.79 | 0.01 |
| **Child sex** | | | | | | | | | |
| Male | 1,003 | 1,105 | 90.8 | Ref. | | | Ref. | | |
| Female | 877 | 950 | 92.3 | 1.23 | 0.89–1.69 | 0.21 | 1.23 | 0.89–1.71 | 0.21 |
| **Age** | | | | | | | | | |
| 0 | 358 | 395 | 90.6 | Ref. | | | Ref. | | |
| 1 | 559 | 611 | 91.5 | 1.15 | 0.73–1.80 | 0.55 | 1.15 | 0.73–1.83 | 0.54 |
| 2 | 438 | 470 | 93.2 | 1.44 | 0.87–2.39 | 0.15 | 1.49 | 0.89–2.49 | 0.13 |
| 3 | 345 | 382 | 90.3 | 0.97 | 0.59–1.58 | 0.89 | 0.89 | 0.53–1.47 | 0.64 |
| 4 | 180 | 197 | 91.4 | 1.14 | 0.62–2.11 | 0.68 | 1.01 | 0.53–1.90 | 0.99 |
| **Caregiver age** | | | | | | | | | |
| <25 | 713 | 791 | 90.1 | Ref. | | | Ref. | | |
| 25–34 | 752 | 818 | 91.9 | 1.25 | 0.88–1.77 | 0.22 | 1.33 | 0.92–1.91 | 0.13 |
| 35–44 | 289 | 309 | 93.5 | 1.60 | 0.95–2.69 | 0.08 | 1.81 | 1.04–3.13 | 0.03 |
| >=45 | 125 | 136 | 91.9 | 1.22 | 0.62–2.39 | 0.56 | 1.22 | 0.60–2.49 | 0.59 |
| Not documented | 1 | 1 | 100.0 | NA | | | NA | | |
| **Caregiver sex** | | | | | | | | | |
| Male | 312 | 341 | 91.5 | Ref. | | | Ref. | | |
| Female | 1,568 | 1,714 | 91.5 | 1.03 | 0.67–1.58 | 0.89 | 1.21 | 0.77–1.92 | 0.41 |
| **Health zone** | | | | | | | | | |
| Ipamu | 928 | 1,017 | 91.2 | Ref. | | | Ref. | | |
| Kenge | 451 | 507 | 89.0 | 0.86 | 0.55–1.34 | 0.49 | 0.65 | 0.41–1.03 | 0.07 |
| Kingandu | 501 | 531 | 94.4 | 1.74 | 1.04–2.92 | 0.03 | 1.19 | 0.67–2.12 | 0.56 |
| **CNS involvement** | | | | | | | | | |
| No | 223 | 246 | 90.7 | Ref. | | | Ref. | | |
| Yes | 1,657 | 1,809 | 91.6 | 1.21 | 0.74–1.96 | 0.45 | 1.13 | 0.66–1.92 | 0.65 |
| **Malaria test** | | | | | | | | | |
| Negative / Not done | 35 | 39 | 89.7 | Ref. | | | Ref. | | |
| Positive | 1,845 | 2,016 | 91.5 | 1.18 | 0.40–3.44 | 0.77 | 0.94 | 0.31–2.81 | 0.91 |
| **Enrolment location** | | | | | | | | | |
| CHW | 1,880 | 2,055 | 91.5 | NA | | | NA | | |
| PHC | 0 | 0 | NA | NA | | | NA | | |
| **Enrolled during rainy season**** | | | | | | | | | |
| No | 680 | 738 | 92.1 | Ref. | | | Ref. | | |
| Yes | 1,200 | 1,317 | 91.1 | 0.92 | 0.65–1.28 | 0.61 | 0.97 | 0.67–1.40 | 0.87 |

| | | | | | | | | | |
|---|---|---|---|---|---|---|---|---|---|
| **Enrolled on a workday** | | | | | | | | | |
| No | 439 | 480 | 91.5 | Ref. | | | Ref. | | |
| Yes | 1,441 | 1,575 | 91.5 | 0.99 | 0.68–1.44 | 0.97 | 1.06 | 0.72–1.54 | 0.78 |
| **Enrolled during Covid-19 pandemic** | | | | | | | | | |
| No | 1,659 | 1,817 | 91.3 | Ref. | | | Ref. | | |
| Yes | 221 | 238 | 92.9 | 1.27 | 0.74–2.17 | 0.39 | 0.97 | 0.53–1.78 | 0.93 |
| **Delay to enrolling provider** | | | | | | | | | |
| 0–1 days | 1,119 | 1,189 | 94.1 | Ref. | | | Ref. | | |
| > 1 day | 755 | 860 | 87.8 | 0.45 | 0.33–0.62 | <0.001 | 0.48 | 0.34–0.68 | <0.001 |
| Not documented | 6 | 6 | 100.0 | NA | | | NA | | |
| **Transport to enrolling provider** | | | | | | | | | |
| By foot / at home | 1,745 | 1,912 | 91.3 | Ref. | | | Ref. | | |
| Vehicle | 135 | 143 | 94.4 | 1.56 | 0.74–3.26 | 0.24 | 1.70 | 0.80–3.60 | 0.17 |
| Not documented | 0 | 0 | NA | NA | | | NA | | |
| **Time to RHF (min)** | | | | | | | | | |
| 0–<15 | 1,168 | 1,277 | 91.5 | Ref. | | | Ref. | | |
| 15–<30 | 605 | 657 | 92.1 | 1.21 | 0.82–1.81 | 0.34 | 1.10 | 0.75–1.63 | 0.61 |
| 30–<60 | 94 | 107 | 87.9 | 0.77 | 0.39–1.53 | 0.46 | 0.61 | 0.30–1.24 | 0.17 |
| > 60 | 0 | 0 | NA | NA | | | NA | | |
| Missing | 13 | 14 | 92.9 | 1.30 | 0.16–10.55 | 0.81 | 1.25 | 0.14–10.82 | 0.84 |
| **Perceived severity** | | | | | | | | | |
| Not fatal | 1,017 | 1,109 | 91.7 | Ref. | | | Ref. | | |
| Fatal | 859 | 942 | 91.2 | 0.96 | 0.70–1.33 | 0.82 | 0.94 | 0.67–1.31 | 0.70 |
| Not documented | 4 | 4 | 100.0 | NA | | | NA | | |
| **Home treatment** | | | | | | | | | |
| No | 1,635 | 1,774 | 92.2 | Ref. | | | Ref. | | |
| Yes | 245 | 281 | 87.2 | 0.59 | 0.40–0.89 | 0.01 | 0.69 | 0.45–1.04 | 0.08 |
| Not documented | 0 | 0 | NA | NA | | | NA | | |

* Danger sign involving the central nervous system (CNS): convulsions, unusually sleepy or unconscious
** April-October
